# Causal effects of gut microbiota on 28-day mortality in patients with sepsis: a Mendelian randomization analysis

**DOI:** 10.1101/2025.10.03.25337304

**Authors:** Weiwei Zhu, Ranran Zhang, Chang Hu, Jianguo Li, Bo Hu

## Abstract

Sepsis, which is characterized by the failure of multiple organ, is the leading cause of death in intensive care units. Extensive research has emphasized a significant link between sepsis and the gut microbiota (GM). However, the causal relationship between these two factors, specifically the impact on 28-day mortality, remains unclear. We conducted a Mendelian randomization (MR) study utilizing genome-wide association study (GWAS) summary statistics. The primary analysis method used to establish the causal relationship was the inverse-variance weighted (IVW) method, along with four other MR methods. Furthermore, we assessed heterogeneity and horizontal pleiotropy using Cochran’s Q test, MR-Egger intercept test, respectively. The IVW method showed that 7 GM, mainly including *class Bacteroidia* [odd ratio (OR) 1.565, 95% confidence interval (CI) 1.161-2.108, *P* = 0.003], genus *Methanobrevibacter* (OR 1.347, 95% CI 1.045-1.736, *P* = 0.022) and others were positively associated with 28-day sepsis mortality. Conversely, genetically predicted 4 GM, including class *Lentisphaeria* (OR 0.751, 95% CI 0.588-0.960, *P* = 0.022), genus *Coprococcus2* (OR 0.537, 95% CI 0.315-0.917, *P* = 0.023), and others were negatively related to 28-day sepsis mortality. Heterogeneity and pleiotropy analyses also confirmed the robustness of our findings (all *P* > 0.05). Our study demonstrated a causal relationship between GM and 28-day mortality in sepsis using MR methods

## INTRODUCTION

Sepsis, a systemic disease, is defined as an infection that involves organ dysfunction (1,2). Despite significant progress in targeting the various biological pathways of sepsis, it remains one of the leading causes of mortality worldwide (3). Recognizing sepsis as a global health priority, the World Health Organization has adopted a resolution to improve the prevention, diagnosis, and management of this condition (4). Interestingly, not all infected patients progress to sepsis, and the underlying mechanisms for this discrepancy are still unknown. Therefore, it is crucial to explore therapeutic or preventive targets against sepsis and strive to improve sepsis outcomes.

The gut microbiota (GM) has been experimentally correlated with several kinds of health problems (5,6), such as an infection with Clostridium difficile (7), hypertension (8), obesity (9,10) and asthma (11) have all been linked to a lack of normal GM function. Since the pathophysiology of sepsis is unclear, there is growing confirmation that disturbance of the GM both predisposes individuals to develop sepsis and has a deleterious effect on sepsis outcomes (12,13). A prospective cohort study from 2019 revealed that newborns with increased bacterial diversity and anaerobic bacterial colonization in their gut microbiome had a protective effect against sepsis (14). Additionally, another study demonstrated a significant dose-response relationship between events leading to microbiome disruption and subsequent hospitalization for severe sepsis (15). Fecal microbiota transplantation (primarily attributable to the expansion of butyrate-producing *Bacteroidetes*) has the potential to reverse the worsening of otherwise fatal sepsis by promoting pathogen clearance through the restoration of host immunity in a way that is reliant upon interferon regulatory factor 3 (16). Despite a significant amount of evidence from observational studies indicating a link between the GM and sepsis, establishing a definitive causal relationship, particularly in terms of sepsis-related mortality, need to be further studied.

Associations reported in observational epidemiological studies are often unreliable estimates of causation and may be influenced by confounding variables, selection bias, measurement errors, and other factors that can distort the findings and lead to incorrect interpretations (17). Mendelian randomization (MR), a genetic technique, offers an opportunity to distinguish causal and non-causal effects between exposure and outcome, without the need for animal studies or randomized controlled trials (18,19). Since genetic variation is essentially passed down randomly from parents to children at conception, many variables that complicate the relationship between exposure and the final result are unable to influence the genetic variants. In a similar vein, genetic variants are typically unaffected by the result and hence by reverse causation (17,20). Recently, MR was employed in a study to validate the causal connections between the GM and blood metabolites (BM). BM like glutamic acid seemed to reduce fecal *Oxalobacter*, while metabolites like alanine, glutamate, and 5-methyltetrahydrofolic acid affected *Proteobacteria* members (21). Long et al. performed two-sample MR to identified 17 strong associations between genetic liability in the GM and cancer (22). Another study successfully utilized the MR method to examine the causal effects between GM and sepsis, it revealed there was a positive correlation between the risk of sepsis and increased abundances of *Clostridiaceae1*, *Alloprevotella*, *LachnospiraceaeND3007 group*, and *Terrisporobacter* (23). However, there is a significant lack of clear research on the relationship between GM and sepsis mortality. In the current study, we aim to use MR to investigate the causal relationship between GM and 28-day sepsis mortality.

## METHODS

### Study Design

The flowchart of the study is presented in **Figure 1**. In summary, the GM was considered as the exposure, while 28-day sepsis mortality was the outcome. The MR method was utilized, using summary statistics obtained from genome-wide association studies (GWAS). For the MR analysis, three assumptions were made in this study: 1) the instrumental variables were strongly associated with the exposure variable; 2) the instrumental variables were independent of confounders that affect both the exposure and outcome; and 3) the instrumental variables affected the outcome through the exposure variable (24, 25).

**Fig. 1.**
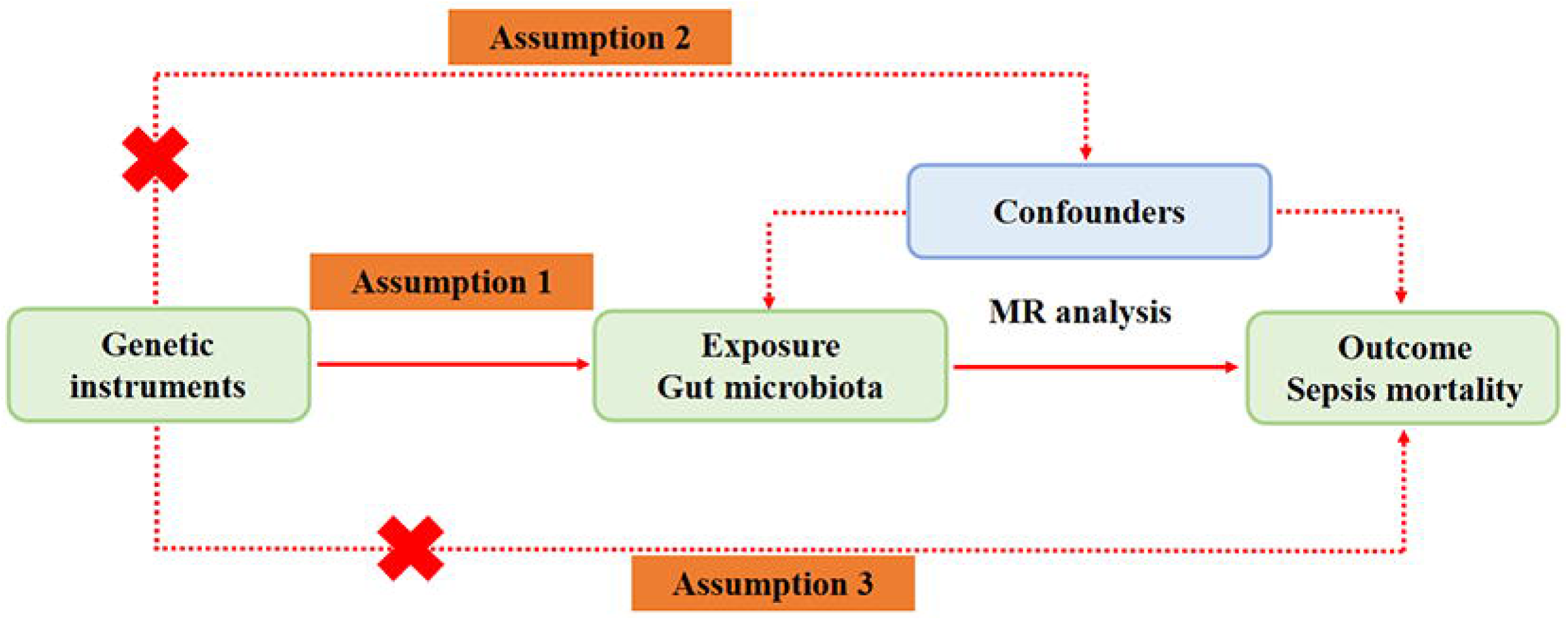
Overall design of the the study. MR, Mendelian randomization.

### Genetic relevant to GM

Genetic association estimates of single nucleotide polymorphisms (SNPs) with GM were obtained from the UK Biobank (26). The UK Biobank is the largest multi-ethnic genome-wide GM meta-analysis conducted to date, analyzing genome-wide genotyping data and 16S fecal microbiota data from 24 cohorts (n=18, 340). The majority of participants had European ancestry (n = 13,266). For profiling the microbial composition and conducting taxonomic classification using direct taxonomic binning, variable regions V4, V3-V4, and V1-V2 of the 16S rRNA gene were targeted. Following processing of the 16S microbiome data, a total of 211 taxa were identified, encompassing 131 genera, 35 families, 20 orders, 16 classes, and 9 phyla (27).

### Genetic relevant to 28-day sepsis mortality

We obtained genetic association estimations for SNPs associated with 28-day sepsis mortality [IEU GWAS ID: ieu-b-5086; N = 486,484 (1896 cases and 484,588 controls)] from the GWAS outcomes of the UK Biobank (28), which gathers detailed phenotypic and genomic information on nearly 500,000 individuals from European ancestry (29,30). The sepsis cases were identified using the International Classification of Diseases (ICD) 10th edition codes A02, A39, A40, and A41, which were obtained from the Integrative Epidemiologic Unit (IEU) GWAS database at https://gwas.mrcieu.ac.uk/ (31–33).

### Instrumental Variables Selection

The instrumental variables (IVs) were selected based on the following criteria (34): (1) IVs were chosen as SNPs associated with each genus at a significance threshold of *P* < 1.0×10^-5^; (2) Linkage disequilibrium (LD) between SNPs was calculated, and only SNPs with the lowest p-value among those with R^2^ < 0.001 (within a clumping window size of 10,000kb) were retained; (3) SNPs with a minor allele frequency (MAF) ≤ 0.01 were removed; (4) For palindromic SNPs, the alleles of the forward strand were determined using allele frequency information. This rigorous selection process aimed to ensure robust instrumental variables for the analysis while considering LD, MAF, and strand information.

### MR Analysis

In the present study, we employed various analytical methods, including random-effects inverse variance weighted (IVW), MR-Egger regression, simple mode, weighted median, and weighted mode (28,35). The IVW method was chosen as the main analysis due to its common usage in MR studies and its ability to yield precise results when all selected SNPs are valid instrumental variables. The remaining four approaches (MR-Egger regression, simple mode, weighted median, and weighted mode) were used as additional methods for MR analysis (36).

### Sensitivity Analyses

In the sensitivity analyses, we employed six methods to assess the robustness of the results: 1) Cochran’s Q test was used to examine the heterogeneity of the IVW method, with a significance level of *P* < 0.05 indicating heterogeneity (37); 2) The MR-Egger intercept test was conducted to estimate genetic pleiotropy, with a significance level of *P* < 0.05 indicating (38); 3) The Steiger filtering method was used to examine the directionality of the results, ensuring their alignment with the hypothesized direction of effect (39); (4) The MR Pleiotropy RESidual Sum and Outlier (MR-PRESSO) global test were applied to assess overall horizontal pleiotropy by comparing the distances of all SNPs and regression lines with the expected distances under the null hypothesis of no horizontal pleiotropy (40); (5) Leave-one-out analysis was used to systematically remove each SNP from the analysis and examining the remaining SNPs to evaluate their individual impact on the results (28). The funnel plot was implemented to evaluate the possibility of horizontal pleiotropy, providing a visual assessment of the symmetry of the data points, which can indicate the presence of potential biases (41); the scatter plot was used to display SNP-outcome connections versus SNP-exposure associations to provide an instantaneous view of the causal-effect estimate for each unique variant (42). Through these sensitivity analyses, the study aimed to assess the robustness of the findings and evaluate potential biases or confounding factors that could impact the results.

### Statistical Analyses

Causal estimates were shown as odds ratios (OR) with corresponding 95% confidence interval (CI). A significance of *P <* 0.05 was considered statistically significant. The scatter plot, leave-one-out plot, funnel plot and all statistical analysis performed in this study were performed using R software (version 4.2.1) with the ‘‘TwoSampleMR’’ package.

## RESULTS

### Characteristics of Genetic Variants

A total of 2,636 SNPs were selected as IVs for the 211 GM taxa, with one taxon from the Clostridiaceae family (ID 1869) lacking a SNP with *P* < 1.0×10^-5^. Consequently, only 210 taxa were included in the subsequent analysis (**Supplementary Table 1**). The *F* statistics for these genetic instruments were all over 10, suggesting the absence of weak instruments. **Supplementary Table 2** provides detailed information on each MR analysis.

### MR Analysis: GM as Exposure, 28-day sepsis mortality as Outcome

The MR analysis estimates from different methods regarding the causal effects of GM on 28-day mortality in sepsis patients are presented in **Table 1**. The IVW results (**Figure. 2**) showed that class *Bacteroidia* [odd ratio (OR) 1.565, 95% confidence interval (CI) 1.161-2.108, *P* = 0.003], genus *Methanobrevibacter* (OR 1.347, 95% CI 1.045-1.736, *P* = 0.022), genus *Ruminococcaceae UCG004* (OR 1.415, 95% CI 1.029-1.946, *P* = 0.033), genus *Ruminococcus torques group* (OR 1.604, 95% CI 1.134-2.269, *P* = 0.008), genus *Sellimonasand* (OR 1.233, 95% CI 1.041-1.459, *P* = 0.015), genus *Terrisporobacter* (OR 1.434, 95% CI 1.061-2.203, *P* = 0.040), and order *Bacteroidales* (OR 1.565, 95% CI 1.161-2.108, *P* = 0.003) were positively associated with 28-day sepsis mortality. Conversely, genetically predicted class *Lentisphaeria* (OR 0.751, 95% CI 0.588-0.960, *P* = 0.022), genus *Coprococcus2* (OR 0.537, 95% CI 0.315-0.917, *P* = 0.023), order *Victivallales* (OR 0.751, 95% CI 0.588-0.960, *P* = 0.022) and phylum *Lentisphaerae* (OR 0.785, 95% CI 0.618-0.999, *P* = 0.049) negatively related to 28-day sepsis mortality. The above findings of simple mode, weighted median, and weighted mode with those obtained through the IVW approach (**Table 1**). However, the MR-Egger test disagreed with the main analysis for some taxa, specifically genus *Ruminococcaceae UCG004* (OR = 1.682, 95% CI: 0.63-0.91, *P* = 3.05) and genus *Coprococcus2* (OR =1.682, 95% CI: 0.76-0.94, *P* = 1.68×10^-3^). It is worth noting that assuming half of IVs to be invalid weakens the statistical power, and the latter test may produce false negatives when the effect was not strong. **Figure 3** showed scatter plots of each MR analysis methods.

**Fig. 2.**
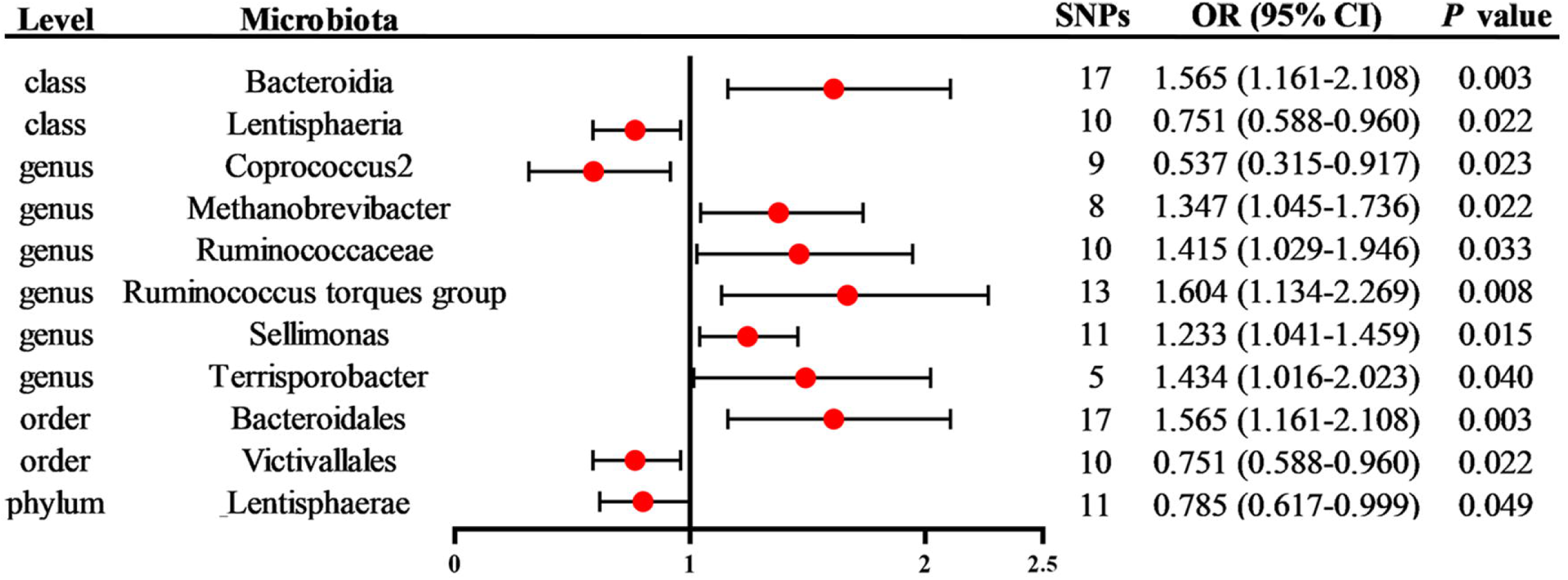
Associations of genetically predicted positive gut microbiota with 28-day sepsis mortality using IVW method. IVW, inverse variance weighted; SNPs, single nucleotide polymorphisms; OR, odds ratio; CI, confidence interval.

**Fig. 3.**
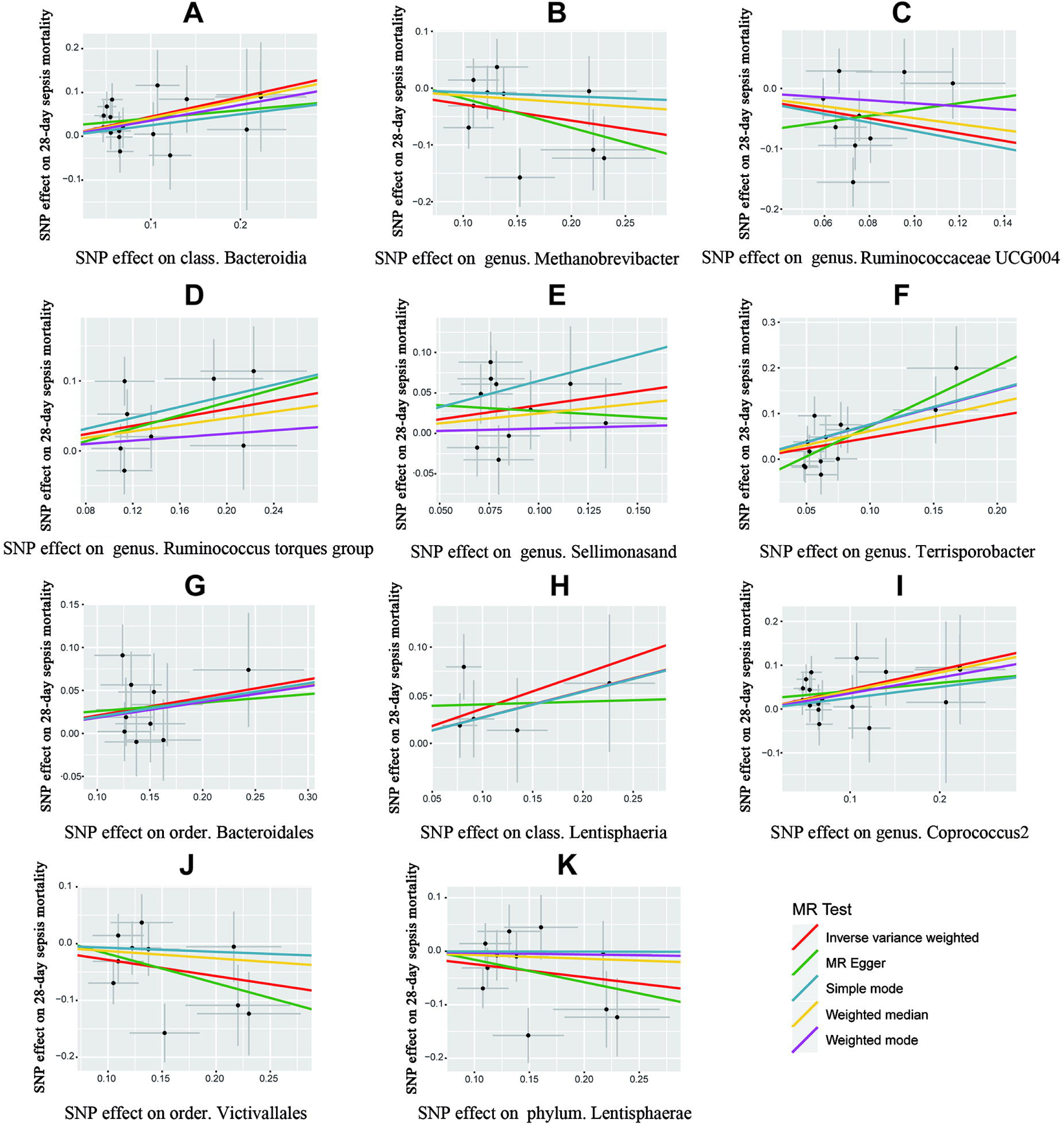
MR scatter plots analysis results of eleven gut microbiotas. Five MR analysis methods (inverse-variance weighted, MR-Egger regression, simple mode, weighted median, and weighted mode) were used to certify the causal effects of GM on 28-day mortality in sepsis, each line represents a different MR method. The genetic link with GM is represented by the X-axis, while the genetic connection with the 28-day sepsis mortality is represented by the Y-axis. (A) *class Bacteroidia;* (B) *genus Methanobrevibacter;* (C) *genus Ruminococcaceae UCG004;* (D) *genus Ruminococcus torques group;* (E) *genus Sellimonasand;* (F) *genus Terrisporobacter; (G) order Bacteroidales;* (H) *class Lentisphaeria;* (I) *genus Coprococcus2;* (J) *order Victivallales;* (K) *phylum Lentisphaerae*. MR, Mendelian randomization; SNP, single nucleotide polymorphism.

**Table 1.** MR estimates for the association between positive gut microbiota and sepsis 28-day mortaliry (*P* < 1 × 10^-5^). MR, Mendelian randomization; SNPs, single nucleotide polymorphisms; Beta, the effect size of the exposure on sepsis 28-day mortaliry; SE, standard error, OR, odds ratio; CI, confidence interval.

| Level | Microbiota | SNPs | Methods | Beta | SE | OR (95% CI) | P value |
| --- | --- | --- | --- | --- | --- | --- | --- |
| class | Bacteroidia | 17 | Inverse variance weighted | 0.448 | 0.152 | 1.565 (1.16-2.11) | 0.003 |
|  |  |  | MR Egger | 0.186 | 0.326 | 1.204 (0.64-2.28) | 0.577 |
|  |  |  | Simple mode | 0.252 | 0.322 | 1.287 (0.68-2.42) | 0.445 |
|  |  |  | Weighted median | 0.416 | 0.217 | 1.516 (0.99-2.32) | 0.055 |
|  |  |  | Weighted mode | 0.358 | 0.288 | 1.431 (0.81-2.52) | 0.231 |
| class | Lentisphaeria | 10 | Inverse variance weighted | -0.286 | 0.125 | 0.751 (0.59-0.96) | 0.022 |
|  |  |  | MR Egger | -0.517 | 0.475 | 0.596 (0.24-1.51) | 0.307 |
|  |  |  | Simple mode | -0.072 | 0.261 | 0.930 (0.56-1.55) | 0.789 |
|  |  |  | Weighted median | -0.130 | 0.152 | 0.878 (0.65-1.18) | 0.393 |
|  |  |  | Weighted mode | -0.072 | 0.241 | 0.930 (0.58-1.49) | 0.771 |
| genus | Coprococcus2 | 9 | Inverse variance weighted | -0.622 | 0.273 | 0.537 (0.31-0.92) | 0.023 |
|  |  |  | MR Egger | 0.520 | 1.540 | 1.682 (0.08-34.41) | 0.746 |
|  |  |  | Simple mode | -0.707 | 0.518 | 0.493 (0.18-1.36) | 0.210 |
|  |  |  | Weighted median | -0.493 | 0.275 | 0.611 (0.36-1.05) | 0.073 |
|  |  |  | Weighted mode | -0.246 | 0.464 | 0.782 (0.31-1.94) | 0.610 |
| genus | Methanobrevibacter | 8 | Inverse variance weighted | 0.298 | 0.130 | 1.347 (1.04-1.74) | 0.022 |
|  |  |  | MR Egger | 0.464 | 0.525 | 1.590 (0.57-4.45) | 0.411 |
|  |  |  | Simple mode | 0.393 | 0.265 | 1.482 (0.88-2.49) | 0.182 |
|  |  |  | Weighted median | 0.233 | 0.154 | 1.262 (0.93-1.71) | 0.131 |
|  |  |  | Weighted mode | 0.122 | 0.257 | 1.130 (0.68-1.87) | 0.650 |
| genus | Ruminococcaceae UCG004 | 10 | Inverse variance weighted | 0.347 | 0.163 | 1.415 (1.03-1.95) | 0.033 |
|  |  |  | MR Egger | -0.143 | 0.879 | 0.866 (0.15-4.86) | 0.875 |
|  |  |  | Simple mode | 0.648 | 0.377 | 1.912 (0.91-4.01) | 0.120 |
|  |  |  | Weighted median | 0.246 | 0.225 | 1.279 (0.82-1.99) | 0.274 |
|  |  |  | Weighted mode | 0.059 | 0.338 | 1.061 (0.55-2.06) | 0.865 |
| genus | Ruminococcus torques group | 13 | Inverse variance weighted | 0.473 | 0.177 | 1.604 (1.13-2.27) | 0.008 |
|  |  |  | MR Egger | 1.324 | 0.523 | 3.757 (1.35-10.47) | 0.028 |
|  |  |  | Simple mode | 0.763 | 0.450 | 2.145 (0.89-5.19) | 0.116 |
|  |  |  | Weighted median | 0.619 | 0.254 | 1.856 (1.13-3.05) | 0.015 |
|  |  |  | Weighted mode | 0.755 | 0.406 | 2.127 (0.96-4.71) | 0.087 |
| genus | Sellimonas | 11 | Inverse variance weighted | 0.209 | 0.086 | 1.233 (1.04-1.46) | 0.015 |
|  |  |  | MR Egger | 0.099 | 0.515 | 1.104 (0.40-3.03) | 0.852 |
|  |  |  | Simple mode | 0.194 | 0.176 | 1.214 (0.86-1.72) | 0.296 |
|  |  |  | Weighted median | 0.186 | 0.115 | 1.205 (0.96-1.51) | 0.105 |
|  |  |  | Weighted mode | 0.183 | 0.172 | 1.201 (0.86-1.68) | 0.313 |
| genus | Terrisporobacter | 5 | Inverse variance weighted | 0.360 | 0.176 | 1.434 (1.02-2.02) | 0.040 |
|  |  |  | MR Egger | 0.029 | 0.481 | 1.029 (0.40-2.64) | 0.956 |
|  |  |  | Simple mode | 0.268 | 0.283 | 1.307 (0.75-2.28) | 0.398 |
|  |  |  | Weighted median | 0.272 | 0.221 | 1.312 (0.85-2.02) | 0.220 |
|  |  |  | Weighted mode | 0.270 | 0.296 | 1.310 (0.73-2.34) | 0.413 |
| order | Bacteroidales | 17 | Inverse variance weighted | 0.448 | 0.152 | 1.565 (1.16-2.11) | 0.003 |
|  |  |  | MR Egger | 0.186 | 0.326 | 1.204 (0.64-2.28) | 0.577 |
|  |  |  | Simple mode | 0.252 | 0.326 | 1.287 (0.68-2.44) | 0.451 |
|  |  |  | Weighted median | 0.416 | 0.224 | 1.516 (0.98-2.35) | 0.063 |
|  |  |  | Weighted mode | 0.358 | 0.296 | 1.431 (0.80-2.56) | 0.244 |
| order | Victivallales | 10 | Inverse variance weighted | -0.286 | 0.125 | 0.751 (0.59-0.96) | 0.022 |
|  |  |  | MR Egger | -0.517 | 0.475 | 0.596 (0.24-1.51) | 0.307 |
|  |  |  | Simple mode | -0.072 | 0.268 | 0.930 (0.55-1.57) | 0.794 |
|  |  |  | Weighted median | -0.130 | 0.152 | 0.878 (0.65-1.18) | 0.391 |
|  |  |  | Weighted mode | -0.072 | 0.241 | 0.930 (0.58-1.49) | 0.772 |
| phylum | Lentisphaerae | 11 | Inverse variance weighted | -0.242 | 0.123 | 0.785 (0.62-1.00) | 0.049 |
|  |  |  | MR Egger | -0.420 | 0.490 | 0.657 (0.25-1.72) | 0.414 |
|  |  |  | Simple mode | -0.003 | 0.243 | 0.997 (0.62-1.61) | 0.990 |
|  |  |  | Weighted median | -0.070 | 0.144 | 0.933 (0.70-1.24) | 0.628 |
|  |  |  | Weighted mode | -0.030 | 0.230 | 0.970 (0.62-1.52) | 0.899 |

### Sensitivity Analysis

Table 2 presents the findings from Cochrane’s Q test, MR-Egger intercept test, Steiger filtering method, and MR-PRESSO global test. The results indicated no potential heterogeneity in the MR analyses of GM and 28-day sepsis mortality. The studies we conducted using the leave-one-out test demonstrated the robustness of the causal effects of GM on the risk of 28-day sepsis mortality (**Figure 4**). There were no noticeable changes observed in the measured causal effects when a single SNP was taken out (all rows were on the same side of 0). Furthermore, the funnel plots showed no apparent horizontal pleiotropy, as the effect size changes symmetrically around the point estimates (**Figure 5**).

**Fig. 4.**
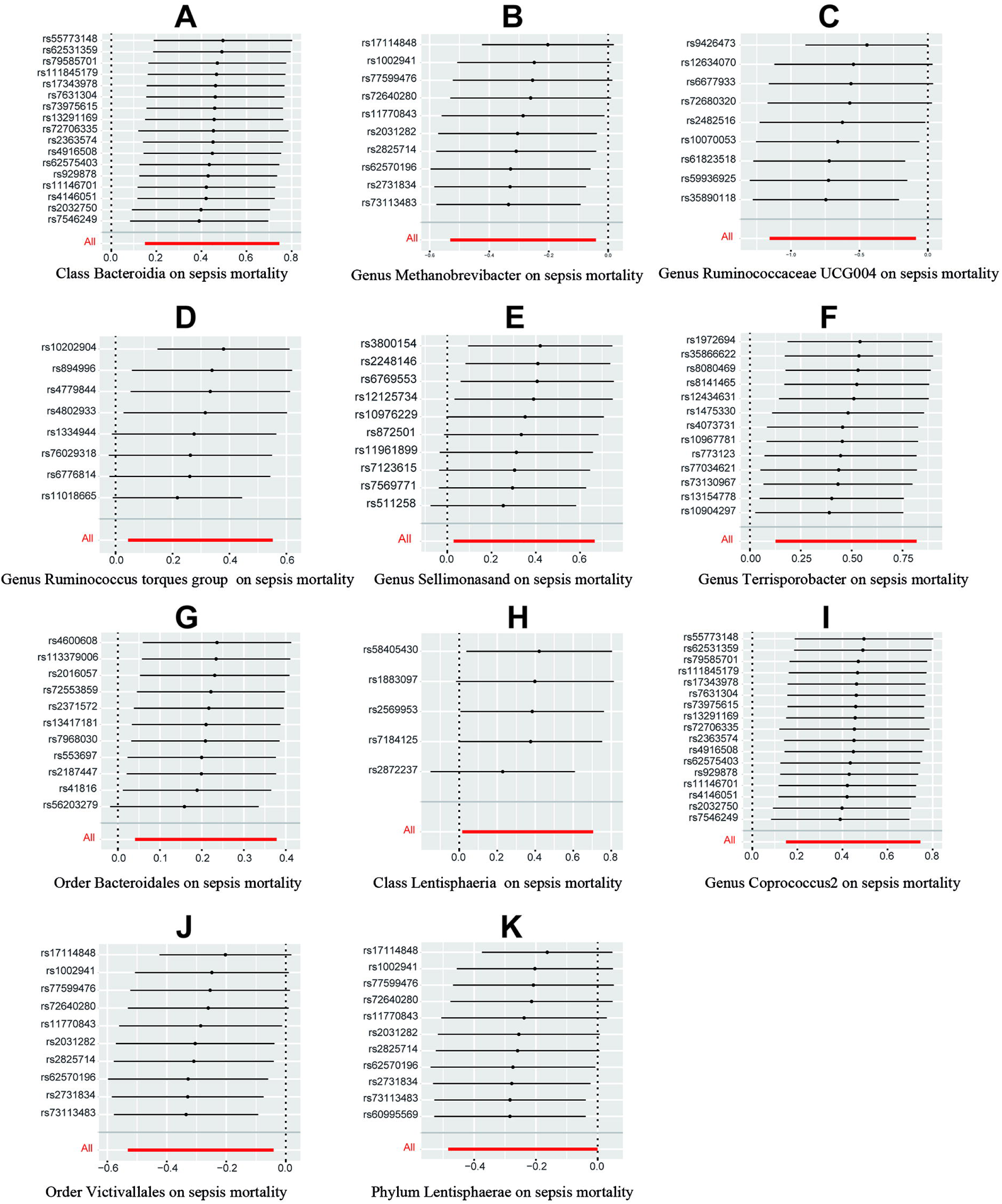
MR leave-one-out sensitivity analysis results of eleven gut microbiotas. Forest plots illustrating the causal effects of 11 gut microbiotas on sepsis mortality by the sequential exclusion of each instrumental variable. When all rows were on the same side of 0, hinting no obvious change by the removal of a single SNP. The beta value and its 95% confidence intervals of causal effects are presented by the horizontal bars. (A) class Bacteroidia; (B) genus Methanobrevibacter; (C) genus Ruminococcaceae UCG004; (D) genus Ruminococcus torques group; (E) genus Sellimonasand; (F) genus Terrisporobacter; (G) order Bacteroidales; (H) class Lentisphaeria; (I) genus Coprococcus2; (J) order Victivallales; (K) phylum Lentisphaerae. Sepsis mortality refers to 28-day mortality. MR, Mendelian randomization; SNP, single nucleotide polymorphism.

**Fig. 5.**
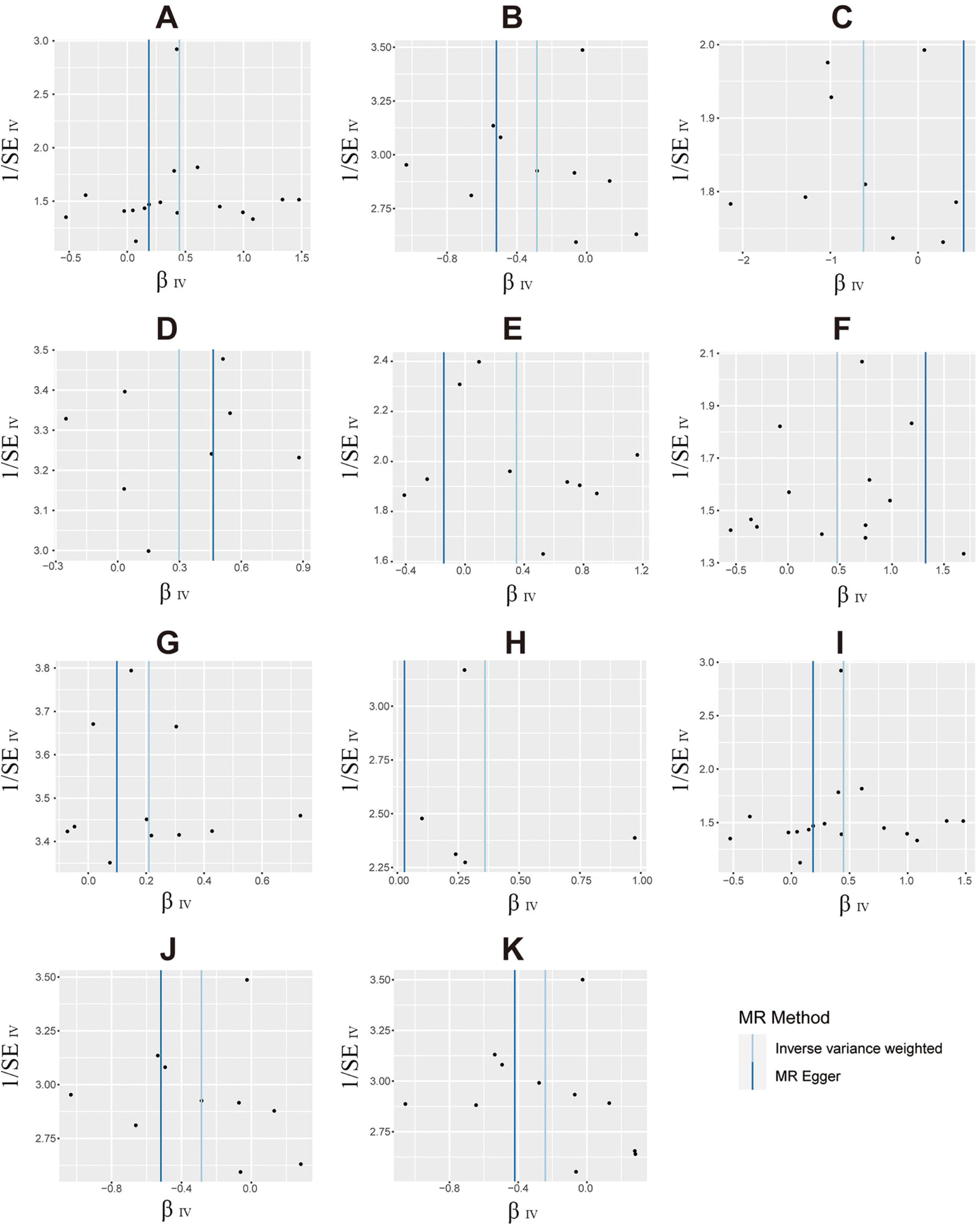
MR funnel plots analysis results of eleven gut microbiotas. The probable existence of horizontal pleiotropy was determined by funnel plots. When the effect size changes symmetrically around the point estimates, hinting no apparent horizontal pleiotropy existed. (A) class Bacteroidia; (B) genus Methanobrevibacter; (C) genus Ruminococcaceae UCG004; (D) genus Ruminococcus torques group; (E) genus Sellimonasand; (F) genus Terrisporobacter; (G) order Bacteroidales; (H) class Lentisphaeria; (I) genus Coprococcus2; (J) order Victivallales; (K) phylum Lentisphaerae. MR, Mendelian randomization; β, the effect size of the exposure on 28-day sepsis mortaliry; SE, standard error; IV, instrumental variable.

**Table 2.** Evaluation of heterogeneity and directional pleiotropy using different methods.

| Level | Microbiota | Cochrane's Q<br>*P* value | MR-Egger |  | Steiger filtering<br>*P* value | MR-PRESSO<br>global *P* value |
| --- | --- | --- | --- | --- | --- | --- |
|  |  |  | Intercept | *P* value |  |  |
| class | Bacteroidia | 0.838 | 0.022 | 0.379 | $3.20 \times 10^{-74}$ | 0.878 |
| class | Lentisphaeria | 0.200 | 0.034 | 0.626 | $2.21 \times 10^{-43}$ | 0.242 |
| genus | Coprococcus2 | 0.020 | -0.087 | 0.476 | $6.79 \times 10^{-42}$ | 0.033 |
| genus | Methanobrevibacter | 0.184 | -0.023 | 0.754 | $2.11 \times 10^{-33}$ | 0.215 |
| genus | Ruminococcaceae | 0.397 | 0.042 | 0.585 | $2.34 \times 10^{-44}$ | 0.417 |
| genus | Ruminococcus torques group | 0.432 | -0.060 | 0.112 | $4.97 \times 10^{-57}$ | 0.466 |
| genus | Sellimonas | 0.768 | 0.016 | 0.832 | $4.53 \times 10^{-49}$ | 0.763 |
| genus | Terrisporobacter | 0.600 | 0.038 | 0.513 | $1.35 \times 10^{-26}$ | 0.684 |
| order | Bacteroidales | 0.838 | 0.022 | 0.379 | $3.20 \times 10^{-74}$ | 0.851 |
| order | Victivallales | 0.200 | 0.034 | 0.626 | $2.21 \times 10^{-43}$ | 0.243 |
| phylum | Lentisphaerae | 0.157 | 0.026 | 0.715 | $1.26 \times 10^{-47}$ | 0.184 |

## DISCUSSION

To the best of our knowledge, this is the first large-scale MR analysis to investigate the causal effects between GM and 28-day sepsis mortality. Our findings indicated that genetically predicted class *Bacteroidia*, genus *Methanobrevibacter*, genus *Ruminococcaceae UCG004*, genus *Ruminococcus torques group*, genus *Sellimonasand*, genus *Terrisporobacter*, and order *Bacteroidales* were positively associated with 28-day sepsis mortality. Conversely, genetically predicted class *Lentisphaeria*, genus *Coprococcus2*, order *Victivallales,* and phylum *Lentisphaerae* were negatively related to 28-day sepsis mortality. These findings underscore the significance role of GM in evaluating 28-day sepsis mortality and indicate that GM may be a target for microbiome-based therapies in sepsis.

There is growing evidence describing the links between GM and sepsis. For instance, a randomized controlled trial conducted by Shimizu et al. (43) demonstrated that prophylactic synbiotics could modulate the abundance of *Bifidobacterium* and *Lactobacillus* in fecal bacteria, potentially preventing enteritis and ventilator-associated pneumonia (VAP) in patients with sepsis. Furthermore, a study with more than 200 preterm newborns discovered that the GM diversity and anaerobic bacterial colonization offer safeguards from sepsis (14). Specifically, within the first weeks of life, decreased *Bacilli* were alongside increased *Gammaproteobacteria, Clostridia, Alphaproteobacteria, Betaproteobacteria*, and *Fusobacteria*. Additionally, a multi-center study conducted in 2020 with 155 patients showed an elevated abundance of microorganisms closely associated with inflammation (such as *Parabacteroides*, *Fusobacterium* and *Bilophila* species) in the microbiota of ICU patients with sepsis (44). During sepsis onset, lower levels of total obligate anaerobes and higher abundance of *Enterococcus* in the GM composition were associated with mortality, although other studies had shown that microbiome diversity does not appear to be associated with mortality in sepsis (45,46). Despite the mounting evidence suggesting that GM affects the risk of sepsis and its related organ injury (30,31), only a limited number of studies in humans have investigated the association between GM features and sepsis mortality, and clinical evidence in many areas is lacking.

MR research can be as easy as testing for a relationship between a SNP and an outcome (47), which allows for whether an exposure influences a medical condition, or as complex as estimating a particular effect using an IV analysis (48, 49). Insomnia (50), differentially expressed genes (51), gut *Prevotellaceae* (35) and synbiotics (43) have been proven to be associated with increased hazard of sepsis with MR analysis.

Different from previous studies, our work specifically investigates the effect of GM on 28-day mortality in sepsis and identified eleven distinct bacterial features that causally affect the risk of sepsis mortality. This study possesses several notable characteristics: 1) unlike conventional observational studies, we employ MR to assess causality of results by utilizing genetic variation. This approach minimizes the potential influence of confounding factors and avoids reverse causality (52); 2) our study benefits from a significantly larger sample size, and the causal associations identified during the initial analysis are subsequently replicated in an independent sample, thereby enhancing confidence in true causality; 3) our methodology leverages existing GWAS studies to screen candidate genetic tool variables as reliable proxies for modifiable exposures. In summary, MR proves to be a robust and effective tool for evaluating the causal relationship between variable and outcome.

The current research does have some limitations, though. Firstly, similar to other studies (53,54), it remains unclear whether these findings can be generalized to the entire popution due to the exclusive inclusion of individuals of European ancestry and the absence of other ethnic groups in the data. Secondly, our study solely relied on genetic-level evidence, which precluded further observational studies and mediating analyses to verify specific regulatory mechanisms for the causal relationship between GM and 28-day sepsis mortality. Thirdly, the breadth of our research was constrained by our MR study’s incapacity to obtain statistics at the person level. Fourth, despite all of MR’s benefits, confounding factors like medication and diet-particularly the use of antibiotics-cannot be totally eliminated. Therefore, further research is necessary to investigate the intricate interplay between GM and sepsis mortality.

## CONCLUSION

In summary, our study supported the casual effect between GM and 28-day sepsis mortality. Specifically, we identified seven bacterial features that displayed a positive causal direction, while four other bacterial traits had a negative causal effect on 28-day sepsis mortality. These findings offer valuable insights that can guide future research endeavors in the field of microbiome-based therapeutics target in sepsis.

## Data Availability

The GM summary data are available in UK Biobank. Sepsis mortality summary data are available at https://gwas.mrcieu.ac.uk/. The GWAS data utilized in this study is publicly available in the public domain, and obtaining individual ethical approval or informed consent was not necessary. The study was conducted in accordance with ethical principles, and the results were reported following the Strengthening the Reporting of Observational Studies in Epidemiology Using Mendelian Randomization (STROBE-MR) guidance from 2021. This guarantees transparency and adherence to best practices in reporting observational studies using MR.

## ACKNOWLEDGEMENTS

Not applicable.

## AUTHOR CONTRIBUTIONS

WZ and BH designed this study; WZ and RZ conducted the manuscript writing and responsible for the data collection; CH was responsible for data analysis; JL and BH critically revising the manuscript. All authors reviewed the manuscript.

## FUNDING

This work was supported by grants from the Key R&D Special Project of Department of Science and Technology of Hubei Province (YFXM2023000244), the Subject Cultivation Project of Zhongnan Hospital of Wuhan University (ZNXKPY2021002), the Health Science and Technology Development Program of Shandong Province (202103031012), the Science and Technology Development Plan of Binzhou Medical University (BY2018KJ18).

## DECLARATIONS

### Ethics approval and consent to participate

Not applicable.

### Consent for publication

Not applicable.

### Competing interests

The authors state that they have no competing interests.

## Notes

### Competing Interest Statement

The authors have declared no competing interest.

