## SUPPLEMENTAL TABLE for "Causal effects of gut microbiota on 28-day mortality in patients with sepsis: a Mendelian randomization analysis"

**Supplemental Table 1 SNPs used as instrumental variables from individual bacterial abundance, the positive gut microbiome and 28-day sepsis mortality (*P* <1.0×10$^{-5}$).**

| Level | Taxa | SNP | effect_allele | other_allele | eaf | beta.exposure | se.exposure | pval.exposure | beta.outcome | se.outcome | pval.outcome | R2 | F |
|---|---|---|---|---|---|---|---|---|---|---|---|---|---|
| class | Bacteroidia id.912 | rs11146701 | A | G | 0.425 | 0.047 | 0.011 | 7.08E-06 | 0.047 | 0.034 | 0.165 | 0.001 | 20.130 |
| class | Bacteroidia id.912 | rs111845179 | T | C | 0.062 | 0.103 | 0.021 | 9.24E-07 | 0.005 | 0.073 | 0.946 | 0.001 | 22.366 |
| class | Bacteroidia id.912 | rs13291169 | C | G | 0.160 | 0.069 | 0.015 | 3.75E-06 | 0.020 | 0.046 | 0.672 | 0.001 | 23.476 |
| class | Bacteroidia id.912 | rs17343978 | A | C | 0.260 | -0.055 | 0.012 | 8.36E-06 | -0.008 | 0.039 | 0.831 | 0.001 | 21.553 |
| class | Bacteroidia id.912 | rs2032750 | C | T | 0.549 | 0.051 | 0.011 | 1.92E-06 | 0.068 | 0.034 | 0.043 | 0.001 | 23.504 |
| class | Bacteroidia id.912 | rs2363574 | T | C | 0.968 | 0.223 | 0.051 | 9.93E-06 | 0.090 | 0.125 | 0.472 | 0.003 | 56.196 |
| class | Bacteroidia id.912 | rs4146051 | G | A | 0.944 | 0.107 | 0.025 | 8.76E-06 | 0.116 | 0.081 | 0.150 | 0.001 | 22.269 |
| class | Bacteroidia id.912 | rs4916508 | A | G | 0.565 | 0.047 | 0.011 | 8.47E-06 | 0.020 | 0.034 | 0.550 | 0.001 | 19.661 |
| class | Bacteroidia id.912 | rs55773148 | G | A | 0.053 | -0.122 | 0.024 | 3.90E-07 | 0.044 | 0.078 | 0.577 | 0.001 | 27.075 |
| class | Bacteroidia id.912 | rs62531359 | T | G | 0.145 | 0.066 | 0.015 | 9.09E-06 | -0.035 | 0.049 | 0.477 | 0.001 | 19.583 |
| class | Bacteroidia id.912 | rs62575403 | C | T | 0.044 | 0.140 | 0.031 | 7.06E-06 | 0.085 | 0.077 | 0.272 | 0.002 | 30.114 |
| class | Bacteroidia id.912 | rs72706335 | T | C | 0.066 | -0.222 | 0.049 | 7.66E-06 | -0.095 | 0.076 | 0.213 | 0.006 | 111.884 |
| class | Bacteroidia id.912 | rs73975615 | G | A | 0.011 | -0.207 | 0.044 | 1.22E-06 | -0.015 | 0.184 | 0.934 | 0.001 | 16.962 |
| class | Bacteroidia id.912 | rs7546249 | A | T | 0.755 | 0.057 | 0.012 | 1.55E-06 | 0.084 | 0.037 | 0.025 | 0.001 | 21.875 |
| class | Bacteroidia id.912 | rs7631304 | G | A | 0.170 | -0.065 | 0.013 | 8.37E-07 | -0.012 | 0.044 | 0.784 | 0.001 | 21.606 |
| class | Bacteroidia id.912 | rs79585701 | A | C | 0.129 | 0.065 | 0.015 | 9.99E-06 | -0.002 | 0.046 | 0.970 | 0.001 | 17.280 |
| class | Bacteroidia id.912 | rs929878 | T | C | 0.756 | 0.055 | 0.012 | 4.73E-06 | 0.044 | 0.038 | 0.249 | 0.001 | 20.407 |
| class | Lentisphaeria id.2250 | rs1002941 | A | G | 0.721 | -0.105 | 0.023 | 8.15E-06 | 0.069 | 0.037 | 0.063 | 0.004 | 81.795 |
| class | Lentisphaeria id.2250 | rs11770843 | C | T | 0.259 | 0.109 | 0.023 | 1.91E-06 | -0.031 | 0.037 | 0.405 | 0.005 | 84.766 |
| class | Lentisphaeria id.2250 | rs17114848 | G | A | 0.109 | 0.152 | 0.032 | 4.06E-06 | -0.157 | 0.052 | 0.002 | 0.005 | 83.280 |
| class | Lentisphaeria id.2250 | rs2031282 | A | G | 0.170 | 0.122 | 0.027 | 4.38E-06 | -0.008 | 0.047 | 0.872 | 0.004 | 77.819 |
| class | Lentisphaeria id.2250 | rs2731834 | G | C | 0.776 | -0.109 | 0.024 | 4.24E-06 | -0.014 | 0.038 | 0.707 | 0.004 | 76.599 |
| class | Lentisphaeria id.2250 | rs2825714 | A | G | 0.157 | -0.137 | 0.029 | 1.72E-06 | 0.010 | 0.047 | 0.836 | 0.005 | 92.161 |
| class | Lentisphaeria id.2250 | rs62570196 | C | T | 0.069 | -0.216 | 0.044 | 1.08E-06 | 0.005 | 0.062 | 0.930 | 0.006 | 110.347 |
| class | Lentisphaeria id.2250 | rs72640280 | A | G | 0.054 | 0.220 | 0.049 | 5.18E-06 | -0.109 | 0.071 | 0.128 | 0.005 | 90.822 |
| class | Lentisphaeria id.2250 | rs73113483 | T | A | 0.123 | -0.131 | 0.029 | 8.66E-06 | -0.037 | 0.050 | 0.456 | 0.004 | 68.517 |
| class | Lentisphaeria id.2250 | rs77599476 | A | G | 0.066 | 0.230 | 0.048 | 1.86E-06 | -0.123 | 0.073 | 0.093 | 0.007 | 120.008 |
| genus | Coprococcus2 id.11302 | rs10070053 | A | G | 0.408 | 0.059 | 0.014 | 7.65E-06 | -0.017 | 0.034 | 0.618 | 0.002 | 31.333 |
| genus | Coprococcus2 id.11302 | rs12634070 | T | C | 0.193 | 0.074 | 0.016 | 9.95E-06 | -0.095 | 0.041 | 0.021 | 0.002 | 31.012 |
| genus | Coprococcus2 id.11302 | rs2482516 | C | T | 0.226 | 0.075 | 0.016 | 4.72E-06 | -0.045 | 0.042 | 0.276 | 0.002 | 36.540 |
| genus | Coprococcus2 id.11302 | rs35890118 | A | G | 0.317 | -0.067 | 0.015 | 8.26E-06 | -0.029 | 0.037 | 0.439 | 0.002 | 35.228 |
| genus | Coprococcus2 id.11302 | rs59936925 | A | T | 0.100 | 0.117 | 0.023 | 9.38E-07 | 0.008 | 0.059 | 0.886 | 0.002 | 45.524 |

| | | | | | | | | | | | | | |
|---|---|---|---|---|---|---|---|---|---|---|---|---|---|
| genus | Coprococcus2 id.11302 | rs61823518 | A | C | 0.095 | -0.096 | 0.022 | 6.68E-06 | -0.027 | 0.055 | 0.622 | 0.002 | 28.935 |
| genus | Coprococcus2 id.11302 | rs6677933 | C | T | 0.242 | -0.080 | 0.016 | 1.19E-06 | 0.083 | 0.041 | 0.042 | 0.002 | 43.587 |
| genus | Coprococcus2 id.11302 | rs72680320 | T | C | 0.414 | -0.065 | 0.014 | 2.27E-06 | 0.064 | 0.034 | 0.056 | 0.002 | 37.585 |
| genus | Coprococcus2 id.11302 | rs9426473 | A | G | 0.279 | 0.073 | 0.016 | 6.31E-06 | -0.155 | 0.041 | 0.000 | 0.002 | 39.141 |
| genus | Methanobrevibacter id.123 | rs10202904 | G | T | 0.533 | 0.113 | 0.024 | 3.09E-06 | -0.028 | 0.034 | 0.404 | 0.006 | 116.926 |
| genus | Methanobrevibacter id.123 | rs11018665 | A | T | 0.341 | 0.113 | 0.025 | 7.03E-06 | 0.099 | 0.035 | 0.004 | 0.006 | 105.871 |
| genus | Methanobrevibacter id.123 | rs1334944 | T | C | 0.285 | 0.115 | 0.026 | 7.61E-06 | 0.053 | 0.036 | 0.140 | 0.005 | 99.781 |
| genus | Methanobrevibacter id.123 | rs4779844 | C | G | 0.676 | -0.110 | 0.025 | 9.28E-06 | -0.003 | 0.035 | 0.921 | 0.005 | 97.088 |
| genus | Methanobrevibacter id.123 | rs4802933 | G | A | 0.822 | 0.136 | 0.031 | 9.74E-06 | 0.020 | 0.045 | 0.653 | 0.005 | 99.203 |
| genus | Methanobrevibacter id.123 | rs6776814 | T | C | 0.090 | -0.189 | 0.042 | 8.05E-06 | -0.103 | 0.057 | 0.068 | 0.006 | 107.335 |
| genus | Methanobrevibacter id.123 | rs76029318 | T | C | 0.065 | 0.223 | 0.045 | 1.08E-06 | 0.114 | 0.064 | 0.076 | 0.006 | 110.725 |
| genus | Methanobrevibacter id.123 | rs894996 | C | A | 0.072 | 0.214 | 0.046 | 3.82E-06 | 0.007 | 0.063 | 0.907 | 0.006 | 112.559 |
| genus | Ruminococcaceae UCG004 id.11362 | rs10976229 | T | G | 0.141 | 0.096 | 0.021 | 7.04E-06 | 0.029 | 0.049 | 0.553 | 0.002 | 41.059 |
| genus | Ruminococcaceae UCG004 id.11362 | rs11961899 | G | A | 0.271 | -0.071 | 0.016 | 9.18E-06 | -0.049 | 0.037 | 0.186 | 0.002 | 36.408 |
| genus | Ruminococcaceae UCG004 id.11362 | rs12125734 | G | T | 0.087 | 0.134 | 0.026 | 2.09E-07 | 0.013 | 0.056 | 0.821 | 0.003 | 52.164 |
| genus | Ruminococcaceae UCG004 id.11362 | rs2248146 | T | C | 0.322 | 0.069 | 0.015 | 8.20E-06 | -0.018 | 0.036 | 0.622 | 0.002 | 38.146 |
| genus | Ruminococcaceae UCG004 id.11362 | rs3800154 | A | C | 0.221 | -0.080 | 0.018 | 6.12E-06 | 0.033 | 0.043 | 0.445 | 0.002 | 40.253 |
| genus | Ruminococcaceae UCG004 id.11362 | rs511258 | G | A | 0.263 | -0.076 | 0.016 | 4.52E-06 | -0.088 | 0.037 | 0.018 | 0.002 | 40.920 |
| genus | Ruminococcaceae UCG004 id.11362 | rs6769553 | A | G | 0.284 | 0.085 | 0.016 | 7.91E-08 | -0.003 | 0.037 | 0.933 | 0.003 | 54.043 |
| genus | Ruminococcaceae UCG004 id.11362 | rs7123615 | C | G | 0.169 | -0.079 | 0.018 | 7.09E-06 | -0.061 | 0.041 | 0.140 | 0.002 | 31.889 |
| genus | Ruminococcaceae UCG004 id.11362 | rs7569771 | A | G | 0.217 | -0.076 | 0.017 | 8.12E-06 | -0.068 | 0.041 | 0.096 | 0.002 | 35.947 |
| genus | Ruminococcaceae UCG004 id.11362 | rs872501 | G | A | 0.127 | 0.116 | 0.026 | 5.81E-06 | 0.061 | 0.071 | 0.390 | 0.003 | 55.073 |
| genus | Ruminococcus torques group id.14377 | rs10904297 | A | G | 0.030 | -0.168 | 0.039 | 2.69E-06 | -0.200 | 0.092 | 0.029 | 0.002 | 29.907 |
| genus | Ruminococcus torques group id.14377 | rs10967781 | C | A | 0.318 | 0.051 | 0.011 | 8.37E-06 | 0.038 | 0.035 | 0.281 | 0.001 | 20.553 |
| genus | Ruminococcus torques group id.14377 | rs12434631 | A | G | 0.140 | 0.075 | 0.015 | 2.77E-06 | 0.001 | 0.048 | 0.988 | 0.001 | 24.701 |
| genus | Ruminococcus torques group id.14377 | rs13154778 | T | A | 0.186 | 0.056 | 0.013 | 7.16E-06 | 0.095 | 0.042 | 0.024 | 0.001 | 17.633 |
| genus | Ruminococcus torques group id.14377 | rs1475330 | C | T | 0.699 | -0.052 | 0.012 | 8.13E-06 | -0.017 | 0.037 | 0.646 | 0.001 | 21.179 |
| genus | Ruminococcus torques group id.14377 | rs1972694 | T | A | 0.184 | -0.061 | 0.014 | 8.93E-06 | 0.034 | 0.043 | 0.432 | 0.001 | 20.781 |
| genus | Ruminococcus torques group id.14377 | rs35866622 | T | C | 0.440 | -0.061 | 0.011 | 2.21E-08 | 0.005 | 0.034 | 0.888 | 0.002 | 33.919 |
| genus | Ruminococcus torques group id.14377 | rs4073731 | T | C | 0.176 | 0.065 | 0.014 | 4.05E-06 | 0.049 | 0.047 | 0.299 | 0.001 | 22.625 |
| genus | Ruminococcus torques group id.14377 | rs73130967 | A | T | 0.112 | 0.077 | 0.017 | 3.71E-06 | 0.076 | 0.050 | 0.132 | 0.001 | 21.724 |
| genus | Ruminococcus torques group id.14377 | rs77034621 | T | G | 0.053 | -0.152 | 0.034 | 6.07E-06 | -0.108 | 0.073 | 0.141 | 0.002 | 42.151 |
| genus | Ruminococcus torques group id.14377 | rs773123 | T | A | 0.112 | 0.082 | 0.017 | 1.59E-06 | 0.065 | 0.051 | 0.205 | 0.001 | 24.859 |
| genus | Ruminococcus torques group id.14377 | rs8080469 | G | A | 0.519 | 0.049 | 0.011 | 3.50E-06 | -0.017 | 0.033 | 0.602 | 0.001 | 22.075 |
| genus | Ruminococcus torques group id.14377 | rs8141465 | G | A | 0.457 | -0.048 | 0.011 | 9.65E-06 | 0.014 | 0.033 | 0.668 | 0.001 | 21.064 |
| genus | Sellimonas id.14369 | rs113379006 | T | C | 0.168 | -0.163 | 0.036 | 7.21E-06 | 0.008 | 0.047 | 0.870 | 0.007 | 136.815 |

| | | | | | | | | | | | | | |
|---|---|---|---|---|---|---|---|---|---|---|---|---|---|
| genus | Sellimonas id.14369 | rs13417181 | T | C | 0.216 | 0.167 | 0.034 | 7.62E-07 | 0.034 | 0.048 | 0.487 | 0.009 | 173.653 |
| genus | Sellimonas id.14369 | rs2016057 | C | A | 0.609 | 0.126 | 0.026 | 1.03E-06 | 0.002 | 0.034 | 0.950 | 0.008 | 139.394 |
| genus | Sellimonas id.14369 | rs2187447 | A | C | 0.061 | 0.243 | 0.053 | 3.98E-06 | 0.074 | 0.066 | 0.266 | 0.007 | 124.592 |
| genus | Sellimonas id.14369 | rs2371572 | A | C | 0.549 | 0.127 | 0.025 | 4.46E-07 | 0.019 | 0.034 | 0.574 | 0.008 | 148.483 |
| genus | Sellimonas id.14369 | rs41816 | A | G | 0.252 | 0.132 | 0.029 | 8.39E-06 | 0.057 | 0.039 | 0.143 | 0.007 | 121.464 |
| genus | Sellimonas id.14369 | rs4600608 | G | A | 0.761 | 0.137 | 0.030 | 4.95E-06 | -0.010 | 0.040 | 0.805 | 0.007 | 126.197 |
| genus | Sellimonas id.14369 | rs553697 | C | T | 0.838 | 0.154 | 0.034 | 6.13E-06 | 0.048 | 0.045 | 0.284 | 0.006 | 118.485 |
| genus | Sellimonas id.14369 | rs56203279 | T | C | 0.330 | -0.124 | 0.027 | 3.72E-06 | -0.091 | 0.036 | 0.011 | 0.007 | 125.630 |
| genus | Sellimonas id.14369 | rs72553859 | G | C | 0.175 | -0.150 | 0.033 | 5.38E-06 | -0.011 | 0.045 | 0.802 | 0.007 | 120.657 |
| genus | Sellimonas id.14369 | rs7968030 | A | T | 0.283 | -0.127 | 0.028 | 5.56E-06 | -0.028 | 0.037 | 0.457 | 0.007 | 121.111 |
| genus | Terrisporobacter id.11348 | rs1883097 | C | T | 0.056 | 0.226 | 0.045 | 4.16E-07 | 0.062 | 0.071 | 0.382 | 0.005 | 99.413 |
| genus | Terrisporobacter id.11348 | rs2569953 | C | A | 0.581 | 0.078 | 0.017 | 8.95E-06 | 0.019 | 0.034 | 0.580 | 0.003 | 53.884 |
| genus | Terrisporobacter id.11348 | rs2872237 | A | C | 0.566 | 0.081 | 0.018 | 3.97E-06 | 0.079 | 0.034 | 0.020 | 0.003 | 59.977 |
| genus | Terrisporobacter id.11348 | rs58405430 | G | T | 0.115 | 0.135 | 0.030 | 7.94E-06 | 0.014 | 0.054 | 0.802 | 0.004 | 68.048 |
| genus | Terrisporobacter id.11348 | rs7184125 | T | C | 0.233 | 0.091 | 0.021 | 8.48E-06 | 0.025 | 0.040 | 0.527 | 0.003 | 54.680 |
| order | Bacteroidales id.913 | rs11146701 | A | G | 0.425 | 0.047 | 0.011 | 7.08E-06 | 0.047 | 0.034 | 0.165 | 0.001 | 20.130 |
| order | Bacteroidales id.913 | rs111845179 | T | C | 0.062 | 0.103 | 0.021 | 9.24E-07 | 0.005 | 0.073 | 0.946 | 0.001 | 22.366 |
| order | Bacteroidales id.913 | rs13291169 | C | G | 0.160 | 0.069 | 0.015 | 3.75E-06 | 0.020 | 0.046 | 0.672 | 0.001 | 23.476 |
| order | Bacteroidales id.913 | rs17343978 | A | C | 0.260 | -0.055 | 0.012 | 8.36E-06 | -0.008 | 0.039 | 0.831 | 0.001 | 21.553 |
| order | Bacteroidales id.913 | rs2032750 | C | T | 0.549 | 0.051 | 0.011 | 1.92E-06 | 0.068 | 0.034 | 0.043 | 0.001 | 23.504 |
| order | Bacteroidales id.913 | rs2363574 | T | C | 0.968 | 0.223 | 0.051 | 9.93E-06 | 0.090 | 0.125 | 0.472 | 0.003 | 56.196 |
| order | Bacteroidales id.913 | rs4146051 | G | A | 0.944 | 0.107 | 0.025 | 8.76E-06 | 0.116 | 0.081 | 0.150 | 0.001 | 22.269 |
| order | Bacteroidales id.913 | rs4916508 | A | G | 0.565 | 0.047 | 0.011 | 8.47E-06 | 0.020 | 0.034 | 0.550 | 0.001 | 19.661 |
| order | Bacteroidales id.913 | rs55773148 | G | A | 0.053 | -0.122 | 0.024 | 3.90E-07 | 0.044 | 0.078 | 0.577 | 0.001 | 27.075 |
| order | Bacteroidales id.913 | rs62531359 | T | G | 0.145 | 0.066 | 0.015 | 9.09E-06 | -0.035 | 0.049 | 0.477 | 0.001 | 19.583 |
| order | Bacteroidales id.913 | rs62575403 | C | T | 0.044 | 0.140 | 0.031 | 7.06E-06 | 0.085 | 0.077 | 0.272 | 0.002 | 30.114 |
| order | Bacteroidales id.913 | rs72706335 | T | C | 0.066 | -0.222 | 0.049 | 7.66E-06 | -0.095 | 0.076 | 0.213 | 0.006 | 111.884 |
| order | Bacteroidales id.913 | rs73975615 | G | A | 0.011 | -0.207 | 0.044 | 1.22E-06 | -0.015 | 0.184 | 0.934 | 0.001 | 16.962 |
| order | Bacteroidales id.913 | rs7546249 | A | T | 0.755 | 0.057 | 0.012 | 1.55E-06 | 0.084 | 0.037 | 0.025 | 0.001 | 21.875 |
| order | Bacteroidales id.913 | rs7631304 | G | A | 0.170 | -0.065 | 0.013 | 8.37E-07 | -0.012 | 0.044 | 0.784 | 0.001 | 21.606 |
| order | Bacteroidales id.913 | rs79585701 | A | C | 0.129 | 0.065 | 0.015 | 9.99E-06 | -0.002 | 0.046 | 0.970 | 0.001 | 17.280 |
| order | Bacteroidales id.913 | rs929878 | T | C | 0.756 | 0.055 | 0.012 | 4.73E-06 | 0.044 | 0.038 | 0.249 | 0.001 | 20.407 |
| order | Victivallales id.2254 | rs1002941 | A | G | 0.721 | -0.105 | 0.023 | 8.15E-06 | 0.069 | 0.037 | 0.063 | 0.004 | 81.795 |
| order | Victivallales id.2254 | rs11770843 | C | T | 0.259 | 0.109 | 0.023 | 1.91E-06 | -0.031 | 0.037 | 0.405 | 0.005 | 84.766 |
| order | Victivallales id.2254 | rs17114848 | G | A | 0.109 | 0.152 | 0.032 | 4.06E-06 | -0.157 | 0.052 | 0.002 | 0.005 | 83.280 |
| order | Victivallales id.2254 | rs2031282 | A | G | 0.170 | 0.122 | 0.027 | 4.38E-06 | -0.008 | 0.047 | 0.872 | 0.004 | 77.819 |

| | | | | | | | | | | | | | |
|---|---|---|---|---|---|---|---|---|---|---|---|---|---|
| order | Victivallales id.2254 | rs2731834 | G | C | 0.776 | -0.109 | 0.024 | 4.24E-06 | -0.014 | 0.038 | 0.707 | 0.004 | 76.599 |
| order | Victivallales id.2254 | rs2825714 | A | G | 0.157 | -0.137 | 0.029 | 1.72E-06 | 0.010 | 0.047 | 0.836 | 0.005 | 92.161 |
| order | Victivallales id.2254 | rs62570196 | C | T | 0.069 | -0.216 | 0.044 | 1.08E-06 | 0.005 | 0.062 | 0.930 | 0.006 | 110.347 |
| order | Victivallales id.2254 | rs72640280 | A | G | 0.054 | 0.220 | 0.049 | 5.18E-06 | -0.109 | 0.071 | 0.128 | 0.005 | 90.822 |
| order | Victivallales id.2254 | rs73113483 | T | A | 0.123 | -0.131 | 0.029 | 8.66E-06 | -0.037 | 0.050 | 0.456 | 0.004 | 68.517 |
| order | Victivallales id.2254 | rs77599476 | A | G | 0.066 | 0.230 | 0.048 | 1.86E-06 | -0.123 | 0.073 | 0.093 | 0.007 | 120.008 |
| phylum | Lentisphaerae id.2238 | rs1002941 | A | G | 0.721 | -0.108 | 0.023 | 4.31E-06 | 0.069 | 0.037 | 0.063 | 0.005 | 85.972 |
| phylum | Lentisphaerae id.2238 | rs11770843 | C | T | 0.259 | 0.112 | 0.023 | 1.14E-06 | -0.031 | 0.037 | 0.405 | 0.005 | 88.646 |
| phylum | Lentisphaerae id.2238 | rs17114848 | G | A | 0.109 | 0.149 | 0.032 | 6.77E-06 | -0.157 | 0.052 | 0.002 | 0.004 | 79.584 |
| phylum | Lentisphaerae id.2238 | rs2031282 | A | G | 0.170 | 0.120 | 0.027 | 5.86E-06 | -0.008 | 0.047 | 0.872 | 0.004 | 75.341 |
| phylum | Lentisphaerae id.2238 | rs2731834 | G | C | 0.776 | -0.110 | 0.024 | 3.80E-06 | -0.014 | 0.038 | 0.707 | 0.004 | 77.299 |
| phylum | Lentisphaerae id.2238 | rs2825714 | A | G | 0.157 | -0.138 | 0.029 | 1.50E-06 | 0.010 | 0.047 | 0.836 | 0.005 | 93.265 |
| phylum | Lentisphaerae id.2238 | rs60995569 | T | G | 0.090 | -0.161 | 0.034 | 9.19E-06 | -0.045 | 0.060 | 0.459 | 0.004 | 77.431 |
| phylum | Lentisphaerae id.2238 | rs62570196 | C | T | 0.069 | -0.217 | 0.044 | 9.64E-07 | 0.005 | 0.062 | 0.930 | 0.006 | 111.224 |
| phylum | Lentisphaerae id.2238 | rs72640280 | A | G | 0.054 | 0.220 | 0.049 | 5.19E-06 | -0.109 | 0.071 | 0.128 | 0.005 | 90.804 |
| phylum | Lentisphaerae id.2238 | rs73113483 | T | A | 0.123 | -0.132 | 0.029 | 8.79E-06 | -0.037 | 0.050 | 0.456 | 0.004 | 69.004 |
| phylum | Lentisphaerae id.2238 | rs77599476 | A | G | 0.066 | 0.230 | 0.048 | 1.90E-06 | -0.123 | 0.073 | 0.093 | 0.006 | 119.682 |

SNPs, single nucleotide polymorphisms; Beta, the effect size of the exposure on 28-day sepsis mortaliry; SE, standard error; OR, odds ratio; CI, confidence interval; R2, variance explained by the genetic variants; F, the F statistics of instrumental variables.

**Supplementary Table 2 Positive MR results of gut microbiome and 28-day sepsis mortality.**

| Level | Taxa | SNPs | method | Beta | SE | OR (95% CI) | *P* value |
|---|---|---|---|---|---|---|---|
| class | Actinobacteria id.419 | 18 | Inverse variance weighted | 0.087 | 0.143 | 1.091 (0.825-1.442) | 0.542 |
| | | | MR Egger | 0.449 | 0.438 | 1.566 (0.664-3.697) | 0.321 |
| | | | Simple mode | 0.123 | 0.306 | 1.131 (0.621-2.059) | 0.692 |
| | | | Weighted median | 0.174 | 0.189 | 1.190 (0.823-1.722) | 0.355 |
| | | | Weighted mode | 0.152 | 0.275 | 1.165 (0.680-1.995) | 0.586 |
| | Alphaproteobacteria id.2379 | 9 | Inverse variance weighted | -0.114 | 0.166 | 0.892 (0.644-1.235) | 0.492 |
| | | | MR Egger | 0.322 | 0.670 | 1.380 (0.371-5.132) | 0.646 |
| | | | Simple mode | 0.065 | 0.298 | 1.067 (0.595-1.914) | 0.833 |
| | | | Weighted median | 0.017 | 0.216 | 1.017 (0.666-1.552) | 0.938 |
| | | | Weighted mode | 0.057 | 0.302 | 1.059 (0.586-1.915) | 0.854 |
| | Bacilli id.1673 | 22 | Inverse variance weighted | 0.073 | 0.129 | 1.076 (0.835-1.387) | 0.572 |
| | | | MR Egger | 0.341 | 0.336 | 1.407 (0.728-2.717) | 0.322 |
| | | | Simple mode | 0.097 | 0.345 | 1.102 (0.560-2.167) | 0.781 |
| | | | Weighted median | 0.053 | 0.185 | 1.054 (0.733-1.516) | 0.775 |
| | | | Weighted mode | 0.081 | 0.285 | 1.084 (0.620-1.895) | 0.779 |
| | Bacteroidia id.912 | 17 | Inverse variance weighted | 0.448 | 0.152 | 1.565 (1.161-2.108) | 0.003 |
| | | | MR Egger | 0.186 | 0.326 | 1.204 (0.635-2.283) | 0.577 |
| | | | Simple mode | 0.252 | 0.322 | 1.287 (0.685-2.418) | 0.445 |
| | | | Weighted median | 0.416 | 0.217 | 1.516 (0.991-2.321) | 0.055 |
| | | | Weighted mode | 0.358 | 0.288 | 1.431 (0.814-2.515) | 0.231 |
| | | 15 | Inverse variance weighted | -0.178 | 0.166 | 0.837 (0.604-1.159) | 0.284 |
| | Betaproteobacteria id.2867 | | MR Egger | 0.146 | 0.549 | 1.157 (0.395-3.392) | 0.794 |
| | | | Simple mode | -0.448 | 0.370 | 0.639 (0.309-1.320) | 0.246 |
| | | | Weighted median | -0.286 | 0.209 | 0.751 (0.499-1.131) | 0.170 |
| | | | Weighted mode | -0.438 | 0.359 | 0.645 (0.320-1.303) | 0.242 |
| | | | Inverse variance weighted | 0.015 | 0.149 | 1.016 (0.758-1.360) | 0.917 |
| | Clostridia id.1859 | 18 | MR Egger | 0.107 | 0.359 | 1.113 (0.551-2.248) | 0.769 |
| | | | Simple mode | -0.095 | 0.340 | 0.910 (0.467-1.771) | 0.784 |
| | | | Weighted median | 0.176 | 0.208 | 1.193 (0.794-1.792) | 0.397 |
| | | | Weighted mode | 0.180 | 0.270 | 1.197 (0.705-2.030) | 0.514 |
| | Coriobacteriia id.809 | 15 | Inverse variance weighted | 0.061 | 0.154 | 1.063 (0.786-1.438) | 0.693 |
| | | | MR Egger | 0.379 | 0.354 | 1.460 (0.730-2.920) | 0.304 |
| | | | Simple mode | 0.493 | 0.346 | 1.638 (0.831-3.226) | 0.176 |
| | | | Weighted median | 0.077 | 0.226 | 1.080 (0.693-1.682) | 0.735 |
| | | | Weighted mode | 0.078 | 0.272 | 1.081 (0.635-1.840) | 0.779 |
| | Deltaproteobacteria id.3087 | 14 | Inverse variance weighted | 0.099 | 0.162 | 1.104 (0.804-1.517) | 0.540 |
| | | | MR Egger | -0.053 | 0.485 | 0.948 (0.367-2.452) | 0.914 |
| | | | Simple mode | -0.051 | 0.368 | 0.950 (0.461-1.956) | 0.892 |
| | | | Weighted median | 0.021 | 0.228 | 1.021 (0.653-1.597) | 0.927 |
| | | | Weighted mode | 0.013 | 0.314 | 1.013 (0.548-1.874) | 0.968 |
| | Erysipelotrichia id.2147 | 13 | Inverse variance weighted | -0.142 | 0.187 | 0.867 (0.601-1.251) | 0.447 |
| | | | MR Egger | -1.870 | 0.794 | 0.154 (0.032-0.731) | 0.038 |
| | | | Simple mode | -0.296 | 0.420 | 0.744 (0.326-1.694) | 0.494 |
| | | | Weighted median | -0.321 | 0.264 | 0.725 (0.433-1.216) | 0.223 |
| | | | Weighted mode | -0.289 | 0.425 | 0.749 (0.325-1.724) | 0.510 |
| | Gammaproteobacteria id.3303 | 8 | Inverse variance weighted | 0.209 | 0.304 | 1.232 (0.679-2.237) | 0.492 |
| | | | MR Egger | 0.476 | 1.106 | 1.609 (0.184-14.061) | 0.682 |
| | | | Simple mode | -0.303 | 0.531 | 0.739 (0.261-2.090) | 0.586 |
| | | | Weighted median | 0.103 | 0.323 | 1.108 (0.589-2.087) | 0.750 |

| | | | | | | | |
|---|---|---|---|---|---|---|---|
| | | | Weighted mode | -0.043 | 0.501 | 0.958 (0.359-2.557) | 0.934 |
| | Lentisphaeria id.2250 | 10 | Inverse variance weighted | -0.286 | 0.125 | 0.751 (0.588-0.960) | 0.022 |
| | | | MR Egger | -0.517 | 0.475 | 0.596 (0.235-1.511) | 0.307 |
| | | | Simple mode | -0.072 | 0.261 | 0.930 (0.558-1.551) | 0.789 |
| | | | Weighted median | -0.130 | 0.152 | 0.878 (0.652-1.183) | 0.393 |
| | | | Weighted mode | -0.072 | 0.241 | 0.930 (0.581-1.491) | 0.771 |
| | Melainabacteria id.1589 | 12 | Inverse variance weighted | -0.142 | 0.108 | 0.868 (0.702-1.073) | 0.190 |
| | | | MR Egger | -0.152 | 0.315 | 0.859 (0.464-1.592) | 0.640 |
| | | | Simple mode | -0.023 | 0.248 | 0.977 (0.601-1.588) | 0.927 |
| | | | Weighted median | -0.060 | 0.149 | 0.942 (0.703-1.262) | 0.689 |
| | | | Weighted mode | -0.019 | 0.234 | 0.981 (0.620-1.551) | 0.936 |
| | Methanobacteria id.119 | 12 | Inverse variance weighted | 0.092 | 0.138 | 1.096 (0.836-1.437) | 0.507 |
| | | | MR Egger | 0.210 | 0.562 | 1.233 (0.410-3.710) | 0.717 |
| | | | Simple mode | 0.205 | 0.273 | 1.227 (0.718-2.097) | 0.469 |
| | | | Weighted median | 0.074 | 0.137 | 1.077 (0.823-1.410) | 0.588 |
| | | | Weighted mode | -0.001 | 0.261 | 0.999 (0.599-1.666) | 0.997 |
| | Mollicutes id.3920 | 12 | Inverse variance weighted | 0.270 | 0.151 | 1.310 (0.975-1.762) | 0.074 |
| | | | MR Egger | -0.161 | 0.487 | 0.851 (0.327-2.213) | 0.748 |
| | | | Simple mode | 0.368 | 0.351 | 1.444 (0.726-2.873) | 0.317 |
| | | | Weighted median | 0.308 | 0.203 | 1.361 (0.915-2.025) | 0.129 |
| | | | Weighted mode | 0.326 | 0.315 | 1.386 (0.748-2.569) | 0.322 |
| | Negativicutes id.2164 | 13 | Inverse variance weighted | 0.087 | 0.181 | 1.091 (0.764-1.557) | 0.631 |
| | | | MR Egger | -0.166 | 0.584 | 0.847 (0.270-2.659) | 0.781 |
| | | | Simple mode | -0.208 | 0.410 | 0.812 (0.364-1.814) | 0.621 |
| | | | Weighted median | -0.094 | 0.242 | 0.910 (0.566-1.462) | 0.696 |
| | | | Weighted mode | -0.201 | 0.387 | 0.818 (0.383-1.745) | 0.613 |
| | Verrucomicrobiae id.4029 | 12 | Inverse variance weighted | 0.198 | 0.196 | 1.219 (0.830-1.790) | 0.313 |
| | | | MR Egger | -0.795 | 0.659 | 0.452 (0.124-1.645) | 0.256 |
| | | | Simple mode | 0.183 | 0.437 | 1.201 (0.510-2.828) | 0.684 |
| | | | Weighted median | 0.219 | 0.225 | 1.245 (0.802-1.934) | 0.328 |
| | | | Weighted mode | 0.148 | 0.391 | 1.159 (0.539-2.494) | 0.713 |
| family | Acidaminococcaceae id.2166 | 8 | Inverse variance weighted | 0.088 | 0.190 | 1.092 (0.753-1.583) | 0.644 |
| | | | MR Egger | 0.070 | 0.561 | 1.072 (0.357-3.221) | 0.905 |
| | | | Simple mode | 0.270 | 0.395 | 1.310 (0.604-2.840) | 0.517 |
| | | | Weighted median | 0.143 | 0.246 | 1.154 (0.712-1.869) | 0.561 |
| | | | Weighted mode | 0.270 | 0.354 | 1.310 (0.654-2.623) | 0.472 |
| | Actinomycetaceae id.421 | 5 | Inverse variance weighted | 0.183 | 0.184 | 1.201 (0.837-1.724) | 0.320 |
| | | | MR Egger | 0.151 | 0.472 | 1.163 (0.461-2.936) | 0.770 |
| | | | Simple mode | 0.234 | 0.305 | 1.264 (0.695-2.297) | 0.486 |
| | | | Weighted median | 0.240 | 0.219 | 1.272 (0.827-1.955) | 0.273 |
| | | | Weighted mode | 0.264 | 0.314 | 1.302 (0.704-2.411) | 0.448 |
| | Alcaligenaceae id.2875 | 17 | Inverse variance weighted | -0.052 | 0.156 | 0.949 (0.699-1.289) | 0.739 |
| | | | MR Egger | 0.111 | 0.623 | 1.117 (0.329-3.788) | 0.861 |
| | | | Simple mode | -0.222 | 0.393 | 0.801 (0.371-1.729) | 0.580 |
| | | | Weighted median | -0.059 | 0.211 | 0.942 (0.623-1.425) | 0.779 |
| | | | Weighted mode | -0.246 | 0.378 | 0.782 (0.373-1.640) | 0.524 |
| | Bacteroidaceae id.917 | 12 | Inverse variance weighted | -0.203 | 0.192 | 0.816 (0.561-1.188) | 0.289 |
| | | | MR Egger | 0.448 | 1.016 | 1.565 (0.214-11.469) | 0.669 |
| | | | Simple mode | 0.255 | 0.483 | 1.291 (0.501-3.329) | 0.608 |
| | | | Weighted median | -0.128 | 0.253 | 0.880 (0.536-1.443) | 0.611 |
| | | | Weighted mode | -0.697 | 0.477 | 0.498 (0.196-1.268) | 0.172 |
| | Bacteroidales S24 7group | 10 | Inverse variance weighted | 0.161 | 0.206 | 1.175 (0.785-1.759) | 0.433 |

| | | | | | | |
|---|---|---|---|---|---|---|
| id.11173 | | | | | | |
| | | MR Egger | 0.454 | 1.072 | 1.575 (0.193-12.880) | 0.683 |
| | | Simple mode | 0.697 | 0.434 | 2.007 (0.858-4.696) | 0.143 |
| | | Weighted median | 0.184 | 0.222 | 1.203 (0.779-1.858) | 0.406 |
| | | Weighted mode | 0.621 | 0.454 | 1.861 (0.764-4.532) | 0.204 |
| Bifidobacteriaceae id.433 | 17 | Inverse variance weighted | 0.072 | 0.147 | 1.075 (0.805-1.435) | 0.624 |
| | | MR Egger | 0.735 | 0.547 | 2.085 (0.713-6.098) | 0.199 |
| | | Simple mode | 0.354 | 0.387 | 1.425 (0.668-3.041) | 0.373 |
| | | Weighted median | 0.188 | 0.216 | 1.207 (0.790-1.845) | 0.384 |
| | | Weighted mode | 0.224 | 0.298 | 1.251 (0.698-2.244) | 0.463 |
| Clostridiaceae1 id.1869 | 11 | Inverse variance weighted | 0.271 | 0.177 | 1.311 (0.927-1.854) | 0.125 |
| | | MR Egger | 0.352 | 0.521 | 1.422 (0.513-3.945) | 0.516 |
| | | Simple mode | 0.056 | 0.348 | 1.057 (0.534-2.093) | 0.876 |
| | | Weighted median | 0.239 | 0.232 | 1.271 (0.807-2.001) | 0.301 |
| | | Weighted mode | 0.123 | 0.331 | 1.131 (0.591-2.165) | 0.718 |
| Clostridiales vadin BB60 group id.11286 | 15 | Inverse variance weighted | -0.081 | 0.154 | 0.923 (0.683-1.247) | 0.600 |
| | | MR Egger | -0.051 | 0.449 | 0.950 (0.394-2.290) | 0.911 |
| | | Simple mode | -0.335 | 0.283 | 0.716 (0.411-1.247) | 0.257 |
| | | Weighted median | -0.215 | 0.182 | 0.806 (0.564-1.153) | 0.238 |
| | | Weighted mode | -0.310 | 0.280 | 0.733 (0.424-1.268) | 0.286 |
| Coriobacteriaceae id.811 | 15 | Inverse variance weighted | 0.061 | 0.154 | 1.063 (0.786-1.438) | 0.693 |
| | | MR Egger | 0.379 | 0.354 | 1.460 (0.730-2.920) | 0.304 |
| | | Simple mode | 0.493 | 0.351 | 1.638 (0.823-3.259) | 0.182 |
| | | Weighted median | 0.077 | 0.233 | 1.080 (0.684-1.705) | 0.742 |
| | | Weighted mode | 0.078 | 0.273 | 1.081 (0.633-1.846) | 0.781 |
| Defluviitaleaceae id.1924 | 10 | Inverse variance weighted | 0.114 | 0.141 | 1.120 (0.850-1.476) | 0.419 |
| | | MR Egger | -0.444 | 0.451 | 0.642 (0.265-1.553) | 0.354 |
| | | Simple mode | -0.058 | 0.325 | 0.944 (0.499-1.785) | 0.863 |
| | | Weighted median | 0.042 | 0.187 | 1.042 (0.722-1.505) | 0.824 |
| | | Weighted mode | -0.053 | 0.272 | 0.949 (0.557-1.615) | 0.851 |
| Desulfovibrionaceae id.3169 | 12 | Inverse variance weighted | 0.095 | 0.185 | 1.100 (0.766-1.580) | 0.606 |
| | | MR Egger | -0.082 | 0.520 | 0.921 (0.333-2.551) | 0.878 |
| | | Simple mode | -0.229 | 0.373 | 0.795 (0.383-1.653) | 0.552 |
| | | Weighted median | -0.016 | 0.235 | 0.984 (0.621-1.560) | 0.945 |
| | | Weighted mode | -0.060 | 0.320 | 0.942 (0.503-1.764) | 0.855 |
| Enterobacteriaceae id.3469 | 10 | Inverse variance weighted | -0.016 | 0.213 | 0.984 (0.648-1.495) | 0.940 |
| | | MR Egger | -1.537 | 0.834 | 0.215 (0.042-1.104) | 0.103 |
| | | Simple mode | 0.055 | 0.456 | 1.056 (0.432-2.580) | 0.907 |
| | | Weighted median | 0.055 | 0.269 | 1.057 (0.623-1.791) | 0.838 |
| | | Weighted mode | 0.091 | 0.447 | 1.095 (0.456-2.629) | 0.844 |
| Erysipelotrichaceae id.2149 | 13 | Inverse variance weighted | -0.142 | 0.187 | 0.867 (0.601-1.251) | 0.447 |
| | | MR Egger | -1.870 | 0.794 | 0.154 (0.032-0.731) | 0.038 |
| | | Simple mode | -0.296 | 0.419 | 0.744 (0.327-1.692) | 0.493 |
| | | Weighted median | -0.321 | 0.261 | 0.725 (0.435-1.210) | 0.219 |
| | | Weighted mode | -0.289 | 0.405 | 0.749 (0.339-1.658) | 0.490 |
| Family XI id.1936 | 10 | Inverse variance weighted | 0.076 | 0.102 | 1.078 (0.884-1.316) | 0.457 |
| | | MR Egger | -0.284 | 0.540 | 0.753 (0.261-2.171) | 0.614 |
| | | Simple mode | -0.002 | 0.206 | 0.998 (0.667-1.493) | 0.992 |
| | | Weighted median | 0.012 | 0.122 | 1.012 (0.797-1.285) | 0.920 |
| | | Weighted mode | -0.006 | 0.216 | 0.994 (0.650-1.518) | 0.977 |
| Family XIII id.1957 | 12 | Inverse variance weighted | 0.336 | 0.183 | 1.400 (0.977-2.005) | 0.067 |

| | | | | | | |
|---|---|---|---|---|---|---|
| | | MR Egger | 1.752 | 0.689 | 5.767 (1.494-22.254) | 0.029 |
| | | Simple mode | -0.075 | 0.443 | 0.928 (0.390-2.210) | 0.869 |
| | | Weighted median | 0.096 | 0.253 | 1.101 (0.671-1.807) | 0.704 |
| | | Weighted mode | -0.075 | 0.418 | 0.928 (0.409-2.106) | 0.862 |
| Lachnospiraceae id.1987 | 17 | Inverse variance weighted | 0.062 | 0.154 | 1.064 (0.787-1.439) | 0.685 |
| | | MR Egger | -0.333 | 0.378 | 0.717 (0.341-1.505) | 0.393 |
| | | Simple mode | 0.026 | 0.312 | 1.027 (0.557-1.893) | 0.934 |
| | | Weighted median | 0.075 | 0.209 | 1.078 (0.715-1.624) | 0.720 |
| | | Weighted mode | 0.040 | 0.283 | 1.041 (0.598-1.813) | 0.889 |
| Lactobacillaceae id.1836 | 10 | Inverse variance weighted | 0.059 | 0.134 | 1.061 (0.816-1.380) | 0.659 |
| | | MR Egger | -0.038 | 0.390 | 0.962 (0.449-2.065) | 0.924 |
| | | Simple mode | -0.218 | 0.287 | 0.804 (0.459-1.410) | 0.467 |
| | | Weighted median | -0.104 | 0.172 | 0.901 (0.644-1.262) | 0.545 |
| | | Weighted mode | -0.203 | 0.267 | 0.817 (0.484-1.379) | 0.468 |
| Methanobacteriaceae id.121 | 12 | Inverse variance weighted | 0.092 | 0.138 | 1.096 (0.836-1.437) | 0.507 |
| | | MR Egger | 0.210 | 0.562 | 1.233 (0.410-3.710) | 0.717 |
| | | Simple mode | 0.205 | 0.270 | 1.227 (0.723-2.084) | 0.465 |
| | | Weighted median | 0.074 | 0.136 | 1.077 (0.825-1.407) | 0.584 |
| | | Weighted mode | -0.001 | 0.279 | 0.999 (0.578-1.726) | 0.997 |
| Oxalobacteraceae id.2966 | 15 | Inverse variance weighted | -0.010 | 0.092 | 0.990 (0.826-1.186) | 0.910 |
| | | MR Egger | -0.644 | 0.370 | 0.525 (0.254-1.083) | 0.105 |
| | | Simple mode | 0.168 | 0.201 | 1.183 (0.798-1.752) | 0.417 |
| | | Weighted median | 0.107 | 0.123 | 1.113 (0.875-1.416) | 0.383 |
| | | Weighted mode | 0.168 | 0.203 | 1.183 (0.794-1.762) | 0.423 |
| Pasteurellaceae id.3689 | 17 | Inverse variance weighted | 0.112 | 0.106 | 1.119 (0.909-1.376) | 0.290 |
| | | MR Egger | 0.140 | 0.261 | 1.150 (0.689-1.918) | 0.601 |
| | | Simple mode | 0.299 | 0.247 | 1.349 (0.831-2.190) | 0.243 |
| | | Weighted median | 0.153 | 0.146 | 1.165 (0.874-1.552) | 0.297 |
| | | Weighted mode | 0.279 | 0.228 | 1.321 (0.845-2.067) | 0.240 |
| Peptococcaceae id.2024 | 9 | Inverse variance weighted | 0.046 | 0.217 | 1.047 (0.685-1.601) | 0.832 |
| | | MR Egger | -0.241 | 0.547 | 0.786 (0.269-2.297) | 0.673 |
| | | Simple mode | -0.300 | 0.347 | 0.741 (0.375-1.463) | 0.413 |
| | | Weighted median | -0.211 | 0.214 | 0.810 (0.533-1.231) | 0.324 |
| | | Weighted mode | -0.271 | 0.264 | 0.762 (0.455-1.278) | 0.334 |
| Peptostreptococcaceae id.2042 | 14 | Inverse variance weighted | 0.180 | 0.181 | 1.197 (0.840-1.707) | 0.320 |
| | | MR Egger | 0.055 | 0.409 | 1.056 (0.474-2.353) | 0.896 |
| | | Simple mode | 0.652 | 0.414 | 1.920 (0.853-4.324) | 0.139 |
| | | Weighted median | 0.463 | 0.217 | 1.589 (1.039-2.431) | 0.033 |
| | | Weighted mode | 0.539 | 0.345 | 1.714 (0.872-3.369) | 0.142 |
| Porphyromonadaceae id.943 | 11 | Inverse variance weighted | 0.051 | 0.204 | 1.052 (0.706-1.568) | 0.802 |
| | | MR Egger | -0.413 | 0.901 | 0.662 (0.113-3.868) | 0.658 |
| | | Simple mode | -0.014 | 0.410 | 0.986 (0.441-2.202) | 0.973 |
| | | Weighted median | 0.059 | 0.263 | 1.061 (0.634-1.775) | 0.821 |
| | | Weighted mode | 0.005 | 0.404 | 1.005 (0.455-2.218) | 0.991 |
| Prevotellaceae id.960 | 18 | Inverse variance weighted | -0.155 | 0.184 | 0.857 (0.597-1.229) | 0.401 |
| | | MR Egger | -0.399 | 0.610 | 0.671 (0.203-2.219) | 0.522 |
| | | Simple mode | -0.409 | 0.363 | 0.664 (0.326-1.352) | 0.275 |
| | | Weighted median | -0.287 | 0.207 | 0.751 (0.500-1.128) | 0.167 |
| | | Weighted mode | -0.409 | 0.361 | 0.664 (0.327-1.349) | 0.273 |
| Rhodospirillaceae id.2717 | 17 | Inverse variance weighted | 0.090 | 0.129 | 1.094 (0.850-1.407) | 0.485 |
| | | MR Egger | -0.611 | 0.589 | 0.543 (0.171-1.722) | 0.316 |
| | | Simple mode | 0.170 | 0.302 | 1.185 (0.655-2.143) | 0.582 |

| | | | | | | | |
|---|---|---|---|---|---|---|---|
| | | | Weighted median | 0.071 | 0.160 | 1.073 (0.783-1.470) | 0.660 |
| | | | Weighted mode | 0.148 | 0.303 | 1.159 (0.640-2.098) | 0.632 |
| | Rikenellaceae id.967 | 20 | Inverse variance weighted | -0.043 | 0.154 | 0.958 (0.707-1.296) | 0.779 |
| | | | MR Egger | -0.101 | 0.498 | 0.904 (0.341-2.398) | 0.841 |
| | | | Simple mode | -0.140 | 0.423 | 0.869 (0.379-1.992) | 0.744 |
| | | | Weighted median | -0.063 | 0.207 | 0.939 (0.626-1.410) | 0.762 |
| | | | Weighted mode | -0.246 | 0.384 | 0.782 (0.369-1.658) | 0.529 |
| | Ruminococcaceae id.2050 | 11 | Inverse variance weighted | -0.064 | 0.177 | 0.938 (0.663-1.327) | 0.718 |
| | | | MR Egger | 0.580 | 0.391 | 1.786 (0.830-3.843) | 0.172 |
| | | | Simple mode | -0.437 | 0.411 | 0.646 (0.288-1.447) | 0.313 |
| | | | Weighted median | 0.016 | 0.243 | 1.017 (0.632-1.635) | 0.946 |
| | | | Weighted mode | 0.205 | 0.293 | 1.228 (0.691-2.182) | 0.499 |
| | Streptococcaceae id.1850 | 16 | Inverse variance weighted | 0.210 | 0.156 | 1.234 (0.909-1.676) | 0.178 |
| | | | MR Egger | 1.112 | 0.555 | 3.041 (1.024-9.029) | 0.065 |
| | | | Simple mode | 0.582 | 0.416 | 1.790 (0.791-4.048) | 0.183 |
| | | | Weighted median | 0.348 | 0.218 | 1.416 (0.924-2.170) | 0.110 |
| | | | Weighted mode | 0.605 | 0.360 | 1.832 (0.904-3.710) | 0.113 |
| | unknown family id.1000001214 | 11 | Inverse variance weighted | -0.179 | 0.112 | 0.836 (0.671-1.043) | 0.112 |
| | | | MR Egger | -0.056 | 0.327 | 0.946 (0.498-1.797) | 0.868 |
| | | | Simple mode | -0.446 | 0.275 | 0.640 (0.373-1.098) | 0.136 |
| | | | Weighted median | -0.227 | 0.154 | 0.797 (0.589-1.077) | 0.140 |
| | | | Weighted mode | -0.433 | 0.265 | 0.649 (0.385-1.091) | 0.134 |
| | unknown family id.1000005471 | 14 | Inverse variance weighted | 0.179 | 0.137 | 1.196 (0.915-1.564) | 0.190 |
| | | | MR Egger | 0.107 | 0.420 | 1.113 (0.489-2.534) | 0.803 |
| | | | Simple mode | 0.350 | 0.311 | 1.418 (0.772-2.607) | 0.281 |
| | | | Weighted median | 0.253 | 0.186 | 1.288 (0.894-1.856) | 0.174 |
| | | | Weighted mode | 0.350 | 0.283 | 1.418 (0.814-2.472) | 0.239 |
| | unknown family id.1000006161 | 15 | Inverse variance weighted | -0.037 | 0.092 | 0.964 (0.805-1.154) | 0.687 |
| | | | MR Egger | -0.215 | 0.405 | 0.806 (0.365-1.784) | 0.604 |
| | | | Simple mode | 0.053 | 0.217 | 1.054 (0.689-1.613) | 0.812 |
| | | | Weighted median | 0.006 | 0.121 | 1.006 (0.793-1.276) | 0.959 |
| | | | Weighted mode | 0.053 | 0.197 | 1.054 (0.717-1.549) | 0.793 |
| | Veillonellaceae id.2172 | 21 | Inverse variance weighted | 0.091 | 0.124 | 1.095 (0.859-1.397) | 0.463 |
| | | | MR Egger | 0.060 | 0.281 | 1.062 (0.613-1.841) | 0.832 |
| | | | Simple mode | 0.349 | 0.320 | 1.417 (0.758-2.651) | 0.288 |
| | | | Weighted median | 0.187 | 0.171 | 1.205 (0.862-1.686) | 0.276 |
| | | | Weighted mode | 0.225 | 0.246 | 1.252 (0.773-2.028) | 0.372 |
| | Verrucomicrobiaceae id.4036 | 12 | Inverse variance weighted | 0.198 | 0.196 | 1.219 (0.830-1.791) | 0.313 |
| | | | MR Egger | -0.796 | 0.659 | 0.451 (0.124-1.643) | 0.255 |
| | | | Simple mode | 0.183 | 0.426 | 1.201 (0.521-2.768) | 0.676 |
| | | | Weighted median | 0.220 | 0.243 | 1.246 (0.773-2.006) | 0.366 |
| | | | Weighted mode | 0.148 | 0.408 | 1.159 (0.521-2.580) | 0.724 |
| | Victivallaceae id.2255 | 14 | Inverse variance weighted | 0.002 | 0.080 | 1.002 (0.856-1.172) | 0.980 |
| | | | MR Egger | 0.113 | 0.380 | 1.120 (0.532-2.360) | 0.771 |
| | | | Simple mode | -0.143 | 0.170 | 0.867 (0.621-1.210) | 0.417 |
| | | | Weighted median | -0.038 | 0.111 | 0.963 (0.775-1.196) | 0.733 |
| | | | Weighted mode | -0.103 | 0.177 | 0.902 (0.637-1.277) | 0.571 |
| genus | Actinomyces id.423 | 7 | Inverse variance weighted | 0.275 | 0.157 | 1.317 (0.968-1.791) | 0.079 |
| | | | MR Egger | 0.353 | 0.415 | 1.424 (0.632-3.208) | 0.433 |

| | | | | | | |
|---|---|---|---|---|---|---|
| | | Simple mode | 0.457 | 0.317 | 1.579 (0.848-2.941) | 0.200 |
| | | Weighted median | 0.318 | 0.210 | 1.374 (0.910-2.075) | 0.131 |
| | | Weighted mode | 0.427 | 0.281 | 1.533 (0.885-2.657) | 0.178 |
| Adlercreutzia id.812 | 12 | Inverse variance weighted | -0.084 | 0.135 | 0.920 (0.707-1.197) | 0.534 |
| | | MR Egger | -0.247 | 0.652 | 0.781 (0.218-2.802) | 0.712 |
| | | Simple mode | -0.219 | 0.310 | 0.803 (0.437-1.475) | 0.494 |
| | | Weighted median | -0.135 | 0.177 | 0.874 (0.617-1.237) | 0.446 |
| | | Weighted mode | -0.206 | 0.301 | 0.814 (0.451-1.470) | 0.509 |
| Akkermansia id.4037 | 12 | Inverse variance weighted | 0.197 | 0.196 | 1.218 (0.830-1.789) | 0.314 |
| | | MR Egger | -0.799 | 0.658 | 0.450 (0.124-1.633) | 0.253 |
| | | Simple mode | 0.184 | 0.423 | 1.202 (0.525-2.751) | 0.672 |
| | | Weighted median | 0.219 | 0.229 | 1.245 (0.794-1.952) | 0.339 |
| | | Weighted mode | 0.140 | 0.418 | 1.150 (0.507-2.607) | 0.744 |
| Alistipes id.968 | 15 | Inverse variance weighted | -0.309 | 0.177 | 0.734 (0.519-1.040) | 0.082 |
| | | MR Egger | -0.078 | 0.885 | 0.925 (0.163-5.246) | 0.931 |
| | | Simple mode | -0.584 | 0.449 | 0.558 (0.231-1.345) | 0.215 |
| | | Weighted median | -0.421 | 0.246 | 0.656 (0.405-1.063) | 0.087 |
| | | Weighted mode | -0.609 | 0.469 | 0.544 (0.217-1.364) | 0.215 |
| Allisonella id.2174 | 6 | Inverse variance weighted | -0.006 | 0.114 | 0.994 (0.795-1.244) | 0.960 |
| | | MR Egger | -0.251 | 0.745 | 0.778 (0.181-3.353) | 0.754 |
| | | Simple mode | -0.167 | 0.235 | 0.847 (0.534-1.343) | 0.511 |
| | | Weighted median | -0.040 | 0.158 | 0.960 (0.705-1.309) | 0.798 |
| | | Weighted mode | -0.136 | 0.236 | 0.873 (0.549-1.386) | 0.589 |
| Alloprevotella id.961 | 7 | Inverse variance weighted | -0.008 | 0.115 | 0.992 (0.792-1.243) | 0.946 |
| | | MR Egger | -0.867 | 1.126 | 0.420 (0.046-3.819) | 0.476 |
| | | Simple mode | -0.039 | 0.198 | 0.962 (0.652-1.419) | 0.851 |
| | | Weighted median | -0.047 | 0.148 | 0.954 (0.715-1.275) | 0.753 |
| | | Weighted mode | -0.026 | 0.200 | 0.974 (0.658-1.443) | 0.901 |
| Anaerofilum id.2053 | 11 | Inverse variance weighted | -0.025 | 0.106 | 0.976 (0.792-1.202) | 0.817 |
| | | MR Egger | 0.720 | 0.464 | 2.054 (0.827-5.104) | 0.155 |
| | | Simple mode | -0.168 | 0.234 | 0.846 (0.534-1.338) | 0.491 |
| | | Weighted median | -0.111 | 0.144 | 0.895 (0.675-1.187) | 0.441 |
| | | Weighted mode | -0.170 | 0.226 | 0.843 (0.542-1.313) | 0.468 |
| Anaerostipes id.1991 | 13 | Inverse variance weighted | -0.188 | 0.212 | 0.828 (0.547-1.255) | 0.374 |
| | | MR Egger | -0.343 | 0.597 | 0.709 (0.220-2.284) | 0.577 |
| | | Simple mode | -0.693 | 0.538 | 0.500 (0.174-1.435) | 0.222 |
| | | Weighted median | -0.470 | 0.258 | 0.625 (0.377-1.036) | 0.068 |
| | | Weighted mode | -0.666 | 0.545 | 0.514 (0.176-1.494) | 0.245 |
| Anaerotruncus id.2054 | 15 | Inverse variance weighted | 0.056 | 0.168 | 1.057 (0.760-1.471) | 0.741 |
| | | MR Egger | -0.268 | 0.534 | 0.765 (0.269-2.177) | 0.624 |
| | | Simple mode | -0.351 | 0.425 | 0.704 (0.306-1.619) | 0.422 |
| | | Weighted median | 0.029 | 0.233 | 1.030 (0.652-1.626) | 0.900 |
| | | Weighted mode | -0.358 | 0.433 | 0.699 (0.299-1.632) | 0.422 |
| Bacteroides id.918 | 12 | Inverse variance weighted | -0.203 | 0.192 | 0.816 (0.561-1.188) | 0.289 |
| | | MR Egger | 0.448 | 1.016 | 1.565 (0.214-11.469) | 0.669 |
| | | Simple mode | 0.255 | 0.496 | 1.291 (0.488-3.412) | 0.617 |
| | | Weighted median | -0.128 | 0.244 | 0.880 (0.545-1.419) | 0.599 |
| | | Weighted mode | -0.697 | 0.470 | 0.498 (0.198-1.252) | 0.166 |
| Barnesiella id.944 | 17 | Inverse variance weighted | 0.195 | 0.175 | 1.216 (0.863-1.712) | 0.264 |
| | | MR Egger | -0.336 | 0.634 | 0.715 (0.207-2.475) | 0.604 |
| | | Simple mode | 0.445 | 0.416 | 1.561 (0.691-3.527) | 0.300 |
| | | Weighted median | 0.260 | 0.220 | 1.296 (0.842-1.996) | 0.238 |

| | | | | | | |
|---|---|---|---|---|---|---|
| | | Weighted mode | 0.369 | 0.385 | 1.447 (0.681-3.075) | 0.351 |
| Bifidobacterium id.436 | 14 | Inverse variance weighted | 0.186 | 0.147 | 1.204 (0.903-1.606) | 0.206 |
| | | MR Egger | 0.603 | 0.384 | 1.827 (0.861-3.880) | 0.143 |
| | | Simple mode | 0.581 | 0.360 | 1.788 (0.882-3.624) | 0.131 |
| | | Weighted median | 0.232 | 0.217 | 1.261 (0.824-1.932) | 0.285 |
| | | Weighted mode | 0.404 | 0.291 | 1.498 (0.846-2.652) | 0.189 |
| Bilophila id.3170 | 16 | Inverse variance weighted | 0.163 | 0.206 | 1.178 (0.786-1.764) | 0.428 |
| | | MR Egger | -0.464 | 0.823 | 0.629 (0.125-3.153) | 0.582 |
| | | Simple mode | -0.121 | 0.474 | 0.886 (0.350-2.245) | 0.802 |
| | | Weighted median | 0.193 | 0.228 | 1.213 (0.775-1.896) | 0.398 |
| | | Weighted mode | -0.131 | 0.437 | 0.877 (0.373-2.063) | 0.767 |
| Blautia id.1992 | 2 | Inverse variance weighted | 0.063 | 0.523 | 1.065 (0.383-2.968) | 0.903 |
| Butyricicoccus id.2055 | 8 | Inverse variance weighted | 0.053 | 0.222 | 1.054 (0.682-1.630) | 0.812 |
| | | MR Egger | 0.408 | 0.464 | 1.504 (0.606-3.732) | 0.413 |
| | | Simple mode | -0.364 | 0.470 | 0.695 (0.277-1.744) | 0.463 |
| | | Weighted median | 0.051 | 0.271 | 1.053 (0.618-1.792) | 0.850 |
| | | Weighted mode | 0.148 | 0.302 | 1.159 (0.642-2.095) | 0.639 |
| Butyricimonas id.945 | 17 | Inverse variance weighted | 0.046 | 0.125 | 1.047 (0.820-1.337) | 0.714 |
| | | MR Egger | 0.214 | 0.452 | 1.238 (0.511-3.001) | 0.643 |
| | | Simple mode | -0.055 | 0.330 | 0.947 (0.496-1.807) | 0.870 |
| | | Weighted median | -0.017 | 0.175 | 0.983 (0.698-1.385) | 0.922 |
| | | Weighted mode | -0.055 | 0.313 | 0.947 (0.512-1.749) | 0.864 |
| Butyrivibrio id.1993 | 16 | Inverse variance weighted | 0.012 | 0.074 | 1.012 (0.876-1.170) | 0.868 |
| | | MR Egger | 0.224 | 0.326 | 1.251 (0.661-2.369) | 0.503 |
| | | Simple mode | 0.084 | 0.173 | 1.088 (0.774-1.527) | 0.635 |
| | | Weighted median | 0.042 | 0.095 | 1.043 (0.865-1.258) | 0.657 |
| | | Weighted mode | 0.087 | 0.177 | 1.091 (0.770-1.545) | 0.632 |
| Candidatus Soleaferrea id.11350 | 13 | Inverse variance weighted | -0.026 | 0.110 | 0.974 (0.785-1.209) | 0.814 |
| | | MR Egger | -0.391 | 0.418 | 0.676 (0.298-1.536) | 0.370 |
| | | Simple mode | 0.011 | 0.279 | 1.011 (0.585-1.746) | 0.970 |
| | | Weighted median | -0.006 | 0.144 | 0.994 (0.750-1.318) | 0.968 |
| | | Weighted mode | -0.048 | 0.250 | 0.953 (0.583-1.556) | 0.850 |
| Catenibacterium id.2153 | 5 | Inverse variance weighted | -0.101 | 0.129 | 0.904 (0.702-1.164) | 0.434 |
| | | MR Egger | 0.125 | 1.335 | 1.133 (0.083-15.494) | 0.931 |
| | | Simple mode | -0.089 | 0.221 | 0.915 (0.594-1.411) | 0.708 |
| | | Weighted median | -0.089 | 0.175 | 0.915 (0.649-1.290) | 0.612 |
| | | Weighted mode | -0.091 | 0.224 | 0.913 (0.588-1.417) | 0.706 |
| Christensenellaceae R 7group id.11283 | 11 | Inverse variance weighted | -0.060 | 0.220 | 0.942 (0.612-1.450) | 0.786 |
| | | MR Egger | -1.321 | 0.608 | 0.267 (0.081-0.878) | 0.058 |
| | | Simple mode | -0.005 | 0.413 | 0.995 (0.442-2.237) | 0.990 |
| | | Weighted median | -0.087 | 0.267 | 0.917 (0.544-1.546) | 0.745 |
| | | Weighted mode | -0.030 | 0.391 | 0.971 (0.452-2.087) | 0.941 |
| Clostridium innocuum group id.14397 | 11 | Inverse variance weighted | 0.089 | 0.100 | 1.093 (0.899-1.329) | 0.372 |
| | | MR Egger | -0.012 | 0.544 | 0.988 (0.340-2.870) | 0.983 |
| | | Simple mode | 0.222 | 0.236 | 1.248 (0.786-1.982) | 0.370 |
| | | Weighted median | 0.155 | 0.137 | 1.167 (0.892-1.528) | 0.259 |
| | | Weighted mode | 0.225 | 0.243 | 1.253 (0.779-2.015) | 0.375 |
| Clostridium sensustricto1 id.1873 | 9 | Inverse variance weighted | 0.173 | 0.173 | 1.188 (0.847-1.667) | 0.317 |

| | | | | | | |
|---|---|---|---|---|---|---|
| | | MR Egger | -0.027 | 0.385 | 0.973 (0.458-2.070) | 0.946 |
| | | Simple mode | -0.119 | 0.353 | 0.888 (0.445-1.773) | 0.745 |
| | | Weighted median | -0.026 | 0.234 | 0.975 (0.616-1.543) | 0.912 |
| | | Weighted mode | -0.110 | 0.304 | 0.896 (0.493-1.626) | 0.726 |
| Collinsella id.815 | 10 | Inverse variance weighted | -0.296 | 0.206 | 0.744 (0.497-1.115) | 0.152 |
| | | MR Egger | -0.011 | 0.854 | 0.989 (0.186-5.270) | 0.990 |
| | | Simple mode | -0.736 | 0.442 | 0.479 (0.202-1.138) | 0.130 |
| | | Weighted median | -0.518 | 0.256 | 0.596 (0.361-0.983) | 0.043 |
| | | Weighted mode | -0.729 | 0.414 | 0.483 (0.214-1.087) | 0.112 |
| Coprobacter id.949 | 14 | Inverse variance weighted | 0.224 | 0.124 | 1.251 (0.982-1.594) | 0.070 |
| | | MR Egger | 0.749 | 0.346 | 2.114 (1.072-4.170) | 0.052 |
| | | Simple mode | 0.271 | 0.232 | 1.312 (0.833-2.067) | 0.263 |
| | | Weighted median | 0.210 | 0.147 | 1.234 (0.924-1.647) | 0.155 |
| | | Weighted mode | 0.277 | 0.213 | 1.319 (0.869-2.001) | 0.217 |
| Coprococcus1 id.11301 | 13 | Inverse variance weighted | -0.292 | 0.161 | 0.746 (0.545-1.023) | 0.069 |
| | | MR Egger | -0.800 | 0.390 | 0.449 (0.209-0.964) | 0.065 |
| | | Simple mode | 0.004 | 0.423 | 1.004 (0.438-2.301) | 0.992 |
| | | Weighted median | -0.168 | 0.231 | 0.846 (0.538-1.330) | 0.468 |
| | | Weighted mode | 0.011 | 0.428 | 1.011 (0.436-2.340) | 0.981 |
| Coprococcus2 id.11302 | 9 | Inverse variance weighted | -0.622 | 0.273 | 0.537 (0.315-0.917) | 0.023 |
| | | MR Egger | 0.520 | 1.540 | 1.682 (0.082-34.408) | 0.746 |
| | | Simple mode | -0.707 | 0.518 | 0.493 (0.179-1.362) | 0.210 |
| | | Weighted median | -0.493 | 0.275 | 0.611 (0.356-1.047) | 0.073 |
| | | Weighted mode | -0.246 | 0.464 | 0.782 (0.315-1.941) | 0.610 |
| Coprococcus3 id.11303 | 9 | Inverse variance weighted | 0.054 | 0.205 | 1.056 (0.707-1.577) | 0.791 |
| | | MR Egger | -0.783 | 0.773 | 0.457 (0.101-2.080) | 0.345 |
| | | Simple mode | -0.434 | 0.494 | 0.648 (0.246-1.706) | 0.405 |
| | | Weighted median | -0.031 | 0.278 | 0.970 (0.562-1.673) | 0.912 |
| | | Weighted mode | -0.441 | 0.442 | 0.643 (0.270-1.530) | 0.348 |
| Defluviitaleaceae UCG011 id.11287 | 8 | Inverse variance weighted | 0.108 | 0.152 | 1.114 (0.826-1.501) | 0.479 |
| | | MR Egger | -0.359 | 0.513 | 0.698 (0.256-1.908) | 0.510 |
| | | Simple mode | -0.016 | 0.318 | 0.984 (0.528-1.836) | 0.962 |
| | | Weighted median | 0.042 | 0.200 | 1.043 (0.705-1.543) | 0.833 |
| | | Weighted mode | -0.016 | 0.290 | 0.984 (0.558-1.737) | 0.958 |
| Desulfovibrio id.3173 | 10 | Inverse variance weighted | -0.097 | 0.148 | 0.908 (0.679-1.212) | 0.511 |
| | | MR Egger | -0.091 | 0.462 | 0.913 (0.369-2.256) | 0.848 |
| | | Simple mode | -0.338 | 0.339 | 0.713 (0.367-1.386) | 0.345 |
| | | Weighted median | -0.154 | 0.195 | 0.857 (0.585-1.256) | 0.430 |
| | | Weighted mode | -0.293 | 0.302 | 0.746 (0.412-1.349) | 0.357 |
| Dialister id.2183 | 12 | Inverse variance weighted | -0.148 | 0.179 | 0.863 (0.608-1.225) | 0.410 |
| | | MR Egger | 0.670 | 0.735 | 1.954 (0.463-8.248) | 0.384 |
| | | Simple mode | 0.191 | 0.339 | 1.211 (0.623-2.352) | 0.583 |
| | | Weighted median | 0.053 | 0.222 | 1.054 (0.682-1.630) | 0.813 |
| | | Weighted mode | 0.168 | 0.307 | 1.183 (0.648-2.159) | 0.595 |
| Dorea id.1997 | 11 | Inverse variance weighted | -0.033 | 0.213 | 0.967 (0.637-1.468) | 0.876 |
| | | MR Egger | 0.179 | 0.647 | 1.196 (0.336-4.253) | 0.789 |
| | | Simple mode | -0.561 | 0.497 | 0.571 (0.215-1.511) | 0.285 |
| | | Weighted median | -0.079 | 0.264 | 0.924 (0.551-1.551) | 0.765 |
| | | Weighted mode | -0.488 | 0.484 | 0.614 (0.238-1.583) | 0.336 |
| Eggerthella id.819 | 10 | Inverse variance weighted | 0.050 | 0.144 | 1.052 (0.793-1.395) | 0.726 |
| | | MR Egger | 0.586 | 0.671 | 1.796 (0.482-6.687) | 0.408 |

| | | | | | | |
|---|---|---|---|---|---|---|
| | | Simple mode | 0.267 | 0.295 | 1.305 (0.733-2.326) | 0.389 |
| | | Weighted median | 0.154 | 0.171 | 1.166 (0.834-1.631) | 0.368 |
| | | Weighted mode | 0.237 | 0.291 | 1.267 (0.716-2.244) | 0.437 |
| Eisenbergiella id.11304 | 12 | Inverse variance weighted | 0.093 | 0.154 | 1.098 (0.812-1.484) | 0.545 |
| | | MR Egger | 1.224 | 1.191 | 3.402 (0.330-35.123) | 0.328 |
| | | Simple mode | 0.411 | 0.260 | 1.509 (0.907-2.511) | 0.142 |
| | | Weighted median | 0.281 | 0.161 | 1.325 (0.966-1.817) | 0.081 |
| | | Weighted mode | 0.400 | 0.265 | 1.492 (0.887-2.510) | 0.160 |
| Enterorhabdus id.820 | 8 | Inverse variance weighted | 0.189 | 0.211 | 1.207 (0.799-1.826) | 0.371 |
| | | MR Egger | 0.090 | 0.600 | 1.094 (0.337-3.547) | 0.886 |
| | | Simple mode | 0.009 | 0.323 | 1.010 (0.536-1.900) | 0.977 |
| | | Weighted median | 0.011 | 0.219 | 1.011 (0.659-1.551) | 0.961 |
| | | Weighted mode | -0.010 | 0.252 | 0.990 (0.604-1.623) | 0.971 |
| Erysipelatoclostridium id.11381 | 16 | Inverse variance weighted | -0.127 | 0.123 | 0.881 (0.692-1.120) | 0.301 |
| | | MR Egger | 0.095 | 0.489 | 1.099 (0.421-2.868) | 0.849 |
| | | Simple mode | -0.171 | 0.271 | 0.843 (0.496-1.433) | 0.537 |
| | | Weighted median | -0.128 | 0.168 | 0.880 (0.633-1.224) | 0.448 |
| | | Weighted mode | -0.181 | 0.286 | 0.835 (0.476-1.462) | 0.537 |
| Erysipelotrichaceae UCG003 id.11384 | 1 | Wald ratio | 0.286 | 0.442 | 1.331 (0.560-3.162) | 0.518 |
| Escherichia Shigella id.3504 | 15 | Inverse variance weighted | -0.160 | 0.171 | 0.852 (0.610-1.191) | 0.349 |
| | | MR Egger | 1.203 | 0.401 | 3.329 (1.517-7.306) | 0.010 |
| | | Simple mode | -0.282 | 0.420 | 0.754 (0.331-1.718) | 0.512 |
| | | Weighted median | -0.161 | 0.200 | 0.851 (0.575-1.260) | 0.420 |
| | | Weighted mode | -0.200 | 0.386 | 0.819 (0.384-1.743) | 0.612 |
| Eubacterium brachy group id.11296 | 10 | Inverse variance weighted | 0.023 | 0.137 | 1.023 (0.783-1.337) | 0.868 |
| | | MR Egger | -0.063 | 0.479 | 0.939 (0.367-2.400) | 0.898 |
| | | Simple mode | 0.325 | 0.332 | 1.383 (0.722-2.651) | 0.353 |
| | | Weighted median | 0.093 | 0.141 | 1.098 (0.833-1.446) | 0.507 |
| | | Weighted mode | 0.319 | 0.333 | 1.376 (0.716-2.645) | 0.363 |
| Eubacterium coprostanoligenes group id.11375 | 14 | Inverse variance weighted | 0.025 | 0.179 | 1.025 (0.723-1.455) | 0.889 |
| | | MR Egger | 0.063 | 0.721 | 1.065 (0.259-4.378) | 0.932 |
| | | Simple mode | 0.004 | 0.430 | 1.004 (0.432-2.334) | 0.993 |
| | | Weighted median | 0.049 | 0.241 | 1.050 (0.655-1.683) | 0.839 |
| | | Weighted mode | 0.042 | 0.396 | 1.043 (0.480-2.267) | 0.917 |
| Eubacterium eligens group id.14372 | 10 | Inverse variance weighted | 0.317 | 0.203 | 1.373 (0.922-2.045) | 0.119 |
| | | MR Egger | 0.665 | 0.585 | 1.945 (0.618-6.120) | 0.288 |
| | | Simple mode | -0.097 | 0.368 | 0.908 (0.441-1.867) | 0.798 |
| | | Weighted median | -0.024 | 0.247 | 0.976 (0.601-1.584) | 0.922 |
| | | Weighted mode | -0.097 | 0.342 | 0.908 (0.464-1.775) | 0.783 |
| Eubacterium fissicatena group id.14373 | 9 | Inverse variance weighted | 0.021 | 0.105 | 1.021 (0.831-1.256) | 0.841 |
| | | MR Egger | -0.214 | 0.573 | 0.807 (0.263-2.483) | 0.720 |
| | | Simple mode | -0.190 | 0.238 | 0.827 (0.518-1.319) | 0.448 |
| | | Weighted median | -0.004 | 0.137 | 0.996 (0.761-1.304) | 0.976 |
| | | Weighted mode | -0.179 | 0.244 | 0.836 (0.518-1.349) | 0.484 |
| Eubacterium hallii group id.11338 | 16 | Inverse variance weighted | 0.097 | 0.146 | 1.101 (0.828-1.465) | 0.507 |

| | | | | | | |
|---|---|---|---|---|---|---|
| | | MR Egger | -0.053 | 0.315 | 0.948 (0.511-1.758) | 0.868 |
| | | Simple mode | 0.221 | 0.415 | 1.247 (0.553-2.812) | 0.603 |
| | | Weighted median | 0.037 | 0.210 | 1.038 (0.687-1.568) | 0.859 |
| | | Weighted mode | -0.171 | 0.348 | 0.843 (0.426-1.669) | 0.631 |
| Eubacterium nodatum group id.11297 | 11 | Inverse variance weighted | 0.147 | 0.087 | 1.158 (0.975-1.374) | 0.094 |
| | | MR Egger | -0.101 | 0.384 | 0.904 (0.426-1.920) | 0.799 |
| | | Simple mode | 0.201 | 0.196 | 1.223 (0.833-1.794) | 0.328 |
| | | Weighted median | 0.192 | 0.123 | 1.212 (0.953-1.541) | 0.117 |
| | | Weighted mode | 0.201 | 0.195 | 1.223 (0.834-1.793) | 0.327 |
| Eubacterium oxidoreducens group id.11339 | 5 | Inverse variance weighted | 0.100 | 0.163 | 1.105 (0.803-1.521) | 0.539 |
| | | MR Egger | 0.673 | 0.520 | 1.960 (0.707-5.431) | 0.286 |
| | | Simple mode | 0.169 | 0.262 | 1.184 (0.708-1.980) | 0.555 |
| | | Weighted median | 0.119 | 0.205 | 1.127 (0.754-1.684) | 0.561 |
| | | Weighted mode | 0.232 | 0.293 | 1.261 (0.710-2.238) | 0.473 |
| Eubacterium rectale group id.14374 | 12 | Inverse variance weighted | 0.038 | 0.239 | 1.039 (0.650-1.660) | 0.873 |
| | | MR Egger | -1.211 | 0.600 | 0.298 (0.092-0.965) | 0.071 |
| | | Simple mode | -0.486 | 0.455 | 0.615 (0.252-1.499) | 0.308 |
| | | Weighted median | -0.353 | 0.259 | 0.703 (0.423-1.168) | 0.174 |
| | | Weighted mode | -0.507 | 0.419 | 0.602 (0.265-1.370) | 0.252 |
| Eubacterium ruminantium group id.11340 | 18 | Inverse variance weighted | 0.142 | 0.103 | 1.153 (0.942-1.411) | 0.169 |
| | | MR Egger | 0.555 | 0.364 | 1.742 (0.854-3.556) | 0.147 |
| | | Simple mode | 0.108 | 0.257 | 1.114 (0.673-1.844) | 0.679 |
| | | Weighted median | 0.086 | 0.139 | 1.090 (0.829-1.432) | 0.537 |
| | | Weighted mode | 0.118 | 0.234 | 1.125 (0.711-1.780) | 0.620 |
| Eubacterium ventriosum group id.11341 | 13 | Inverse variance weighted | -0.085 | 0.174 | 0.919 (0.653-1.292) | 0.625 |
| | | MR Egger | -0.258 | 0.705 | 0.772 (0.194-3.074) | 0.721 |
| | | Simple mode | 0.109 | 0.353 | 1.115 (0.558-2.230) | 0.763 |
| | | Weighted median | 0.058 | 0.228 | 1.059 (0.678-1.655) | 0.800 |
| | | Weighted mode | 0.100 | 0.340 | 1.105 (0.567-2.151) | 0.775 |
| Eubacterium xylanophilum group id.14375 | 11 | Inverse variance weighted | 0.102 | 0.199 | 1.107 (0.750-1.635) | 0.609 |
| | | MR Egger | 0.135 | 0.680 | 1.145 (0.302-4.342) | 0.847 |
| | | Simple mode | 0.424 | 0.456 | 1.528 (0.625-3.733) | 0.375 |
| | | Weighted median | 0.165 | 0.246 | 1.180 (0.728-1.912) | 0.503 |
| | | Weighted mode | 0.259 | 0.420 | 1.295 (0.568-2.953) | 0.552 |
| Faecalibacterium id.2057 | 13 | Inverse variance weighted | 0.086 | 0.155 | 1.090 (0.804-1.477) | 0.579 |
| | | MR Egger | 0.062 | 0.322 | 1.064 (0.566-2.000) | 0.851 |
| | | Simple mode | -0.086 | 0.345 | 0.917 (0.467-1.804) | 0.807 |
| | | Weighted median | 0.031 | 0.215 | 1.032 (0.677-1.573) | 0.885 |
| | | Weighted mode | -0.012 | 0.262 | 0.988 (0.591-1.652) | 0.964 |
| Family XIII AD3011 group id.11293 | 15 | Inverse variance weighted | -0.074 | 0.152 | 0.929 (0.690-1.250) | 0.625 |
| | | MR Egger | 0.372 | 0.751 | 1.451 (0.333-6.324) | 0.629 |
| | | Simple mode | -0.412 | 0.362 | 0.662 (0.326-1.345) | 0.273 |
| | | Weighted median | -0.094 | 0.206 | 0.911 (0.608-1.364) | 0.649 |
| | | Weighted mode | -0.419 | 0.354 | 0.658 (0.328-1.317) | 0.257 |
| Family XIII UCG001 id.11294 | 10 | Inverse variance weighted | -0.151 | 0.209 | 0.860 (0.571-1.295) | 0.470 |

| | | | | | | |
|---|---|---|---|---|---|---|
| | | MR Egger | -0.045 | 0.639 | 0.956 (0.273-3.344) | 0.946 |
| | | Simple mode | 0.129 | 0.392 | 1.138 (0.527-2.455) | 0.750 |
| | | Weighted median | 0.128 | 0.246 | 1.137 (0.702-1.841) | 0.602 |
| | | Weighted mode | 0.151 | 0.337 | 1.163 (0.600-2.254) | 0.664 |
| Flavonifractor id.2059 | 8 | Inverse variance weighted | 0.061 | 0.181 | 1.063 (0.746-1.515) | 0.736 |
| | | MR Egger | -0.272 | 0.703 | 0.762 (0.192-3.024) | 0.712 |
| | | Simple mode | 0.247 | 0.389 | 1.280 (0.597-2.745) | 0.546 |
| | | Weighted median | 0.119 | 0.234 | 1.127 (0.712-1.783) | 0.611 |
| | | Weighted mode | 0.258 | 0.396 | 1.294 (0.595-2.813) | 0.536 |
| Fusicatenibacter id.11305 | 18 | Inverse variance weighted | -0.081 | 0.158 | 0.923 (0.676-1.258) | 0.611 |
| | | MR Egger | -0.108 | 0.652 | 0.898 (0.250-3.220) | 0.871 |
| | | Simple mode | -0.444 | 0.446 | 0.642 (0.268-1.538) | 0.334 |
| | | Weighted median | -0.133 | 0.224 | 0.876 (0.564-1.360) | 0.555 |
| | | Weighted mode | -0.426 | 0.419 | 0.653 (0.287-1.485) | 0.324 |
| Gordonibacter id.821 | 14 | Inverse variance weighted | 0.136 | 0.084 | 1.146 (0.972-1.351) | 0.104 |
| | | MR Egger | -0.149 | 0.337 | 0.861 (0.444-1.669) | 0.666 |
| | | Simple mode | 0.027 | 0.202 | 1.028 (0.691-1.528) | 0.895 |
| | | Weighted median | 0.095 | 0.111 | 1.099 (0.884-1.367) | 0.396 |
| | | Weighted mode | 0.018 | 0.188 | 1.018 (0.704-1.473) | 0.924 |
| Haemophilus id.3698 | 13 | Inverse variance weighted | 0.177 | 0.118 | 1.194 (0.948-1.504) | 0.133 |
| | | MR Egger | 0.024 | 0.297 | 1.024 (0.572-1.832) | 0.938 |
| | | Simple mode | 0.373 | 0.246 | 1.452 (0.896-2.354) | 0.156 |
| | | Weighted median | 0.170 | 0.150 | 1.185 (0.883-1.590) | 0.258 |
| | | Weighted mode | 0.185 | 0.233 | 1.204 (0.763-1.900) | 0.441 |
| Holdemanella id.11393 | 14 | Inverse variance weighted | 0.213 | 0.110 | 1.237 (0.997-1.536) | 0.054 |
| | | MR Egger | -0.028 | 0.348 | 0.972 (0.491-1.923) | 0.936 |
| | | Simple mode | 0.208 | 0.244 | 1.232 (0.763-1.989) | 0.409 |
| | | Weighted median | 0.207 | 0.144 | 1.229 (0.928-1.629) | 0.150 |
| | | Weighted mode | 0.211 | 0.227 | 1.235 (0.791-1.927) | 0.370 |
| Holdemania id.2157 | 18 | Inverse variance weighted | 0.015 | 0.107 | 1.015 (0.823-1.251) | 0.892 |
| | | MR Egger | -0.466 | 0.311 | 0.627 (0.341-1.155) | 0.154 |
| | | Simple mode | 0.060 | 0.250 | 1.062 (0.651-1.732) | 0.813 |
| | | Weighted median | 0.026 | 0.148 | 1.026 (0.768-1.372) | 0.861 |
| | | Weighted mode | 0.042 | 0.234 | 1.043 (0.659-1.650) | 0.859 |
| Howardella id.2000 | 10 | Inverse variance weighted | 0.039 | 0.099 | 1.039 (0.856-1.263) | 0.697 |
| | | MR Egger | 0.047 | 0.449 | 1.048 (0.435-2.527) | 0.919 |
| | | Simple mode | -0.065 | 0.204 | 0.937 (0.629-1.397) | 0.758 |
| | | Weighted median | 0.021 | 0.123 | 1.021 (0.803-1.300) | 0.863 |
| | | Weighted mode | -0.076 | 0.193 | 0.927 (0.634-1.354) | 0.703 |
| Hungatella id.11306 | 5 | Inverse variance weighted | 0.034 | 0.147 | 1.035 (0.776-1.380) | 0.815 |
| | | MR Egger | 0.820 | 0.932 | 2.270 (0.365-14.116) | 0.444 |
| | | Simple mode | 0.123 | 0.245 | 1.131 (0.699-1.827) | 0.643 |
| | | Weighted median | 0.107 | 0.186 | 1.113 (0.773-1.603) | 0.565 |
| | | Weighted mode | 0.123 | 0.231 | 1.131 (0.719-1.777) | 0.623 |
| Intestinibacter id.11345 | 14 | Inverse variance weighted | -0.162 | 0.146 | 0.851 (0.639-1.132) | 0.268 |
| | | MR Egger | 0.142 | 0.478 | 1.152 (0.452-2.940) | 0.772 |
| | | Simple mode | -0.080 | 0.328 | 0.923 (0.485-1.757) | 0.812 |
| | | Weighted median | -0.149 | 0.190 | 0.862 (0.594-1.250) | 0.434 |
| | | Weighted mode | -0.085 | 0.347 | 0.919 (0.465-1.815) | 0.811 |
| Intestinimonas id.2062 | 20 | Inverse variance weighted | -0.111 | 0.119 | 0.895 (0.709-1.129) | 0.348 |
| | | MR Egger | -0.143 | 0.307 | 0.867 (0.475-1.581) | 0.646 |
| | | Simple mode | -0.392 | 0.301 | 0.676 (0.375-1.219) | 0.209 |

| | | | | | | |
|---|---|---|---|---|---|---|
| | | Weighted median | -0.249 | 0.164 | 0.780 (0.565-1.076) | 0.130 |
| | | Weighted mode | -0.419 | 0.292 | 0.658 (0.371-1.166) | 0.168 |
| Lachnoclostridium id.11308 | 13 | Inverse variance weighted | -0.023 | 0.241 | 0.977 (0.609-1.567) | 0.923 |
| | | MR Egger | 0.027 | 0.903 | 1.028 (0.175-6.036) | 0.976 |
| | | Simple mode | 0.147 | 0.547 | 1.159 (0.396-3.387) | 0.793 |
| | | Weighted median | 0.061 | 0.290 | 1.063 (0.602-1.876) | 0.833 |
| | | Weighted mode | -0.002 | 0.562 | 0.998 (0.331-3.005) | 0.997 |
| Lachnospiraceae FCS020 group id.11314 | 15 | Inverse variance weighted | -0.235 | 0.182 | 0.791 (0.554-1.129) | 0.196 |
| | | MR Egger | -0.570 | 0.459 | 0.565 (0.230-1.391) | 0.236 |
| | | Simple mode | -0.255 | 0.336 | 0.775 (0.401-1.496) | 0.460 |
| | | Weighted median | -0.321 | 0.197 | 0.725 (0.493-1.066) | 0.102 |
| | | Weighted mode | -0.274 | 0.316 | 0.760 (0.409-1.412) | 0.400 |
| Lachnospiraceae NC2004 group id.11316 | 10 | Inverse variance weighted | 0.121 | 0.128 | 1.128 (0.878-1.450) | 0.346 |
| | | MR Egger | 0.358 | 0.543 | 1.430 (0.493-4.146) | 0.529 |
| | | Simple mode | 0.295 | 0.247 | 1.343 (0.828-2.178) | 0.263 |
| | | Weighted median | 0.215 | 0.165 | 1.240 (0.897-1.713) | 0.192 |
| | | Weighted mode | 0.292 | 0.277 | 1.339 (0.778-2.303) | 0.320 |
| Lachnospiraceae ND3007 group id.11317 | 3 | Inverse variance weighted | 0.037 | 0.369 | 1.037 (0.503-2.138) | 0.921 |
| | | MR Egger | 6.809 | 6.205 | 906.046 (0.005-173.289) | 0.470 |
| | | Simple mode | -0.194 | 0.579 | 0.824 (0.265-2.560) | 0.769 |
| | | Weighted median | -0.103 | 0.471 | 0.902 (0.359-2.268) | 0.826 |
| | | Weighted mode | -0.210 | 0.552 | 0.810 (0.275-2.390) | 0.740 |
| Lachnospiraceae NK4A136 group id.11319 | 16 | Inverse variance weighted | -0.088 | 0.143 | 0.916 (0.692-1.211) | 0.536 |
| | | MR Egger | 0.092 | 0.302 | 1.096 (0.607-1.981) | 0.766 |
| | | Simple mode | -0.164 | 0.324 | 0.849 (0.450-1.601) | 0.619 |
| | | Weighted median | -0.139 | 0.195 | 0.870 (0.593-1.276) | 0.476 |
| | | Weighted mode | -0.020 | 0.280 | 0.980 (0.566-1.697) | 0.943 |
| Lachnospiraceae UCG001 id.11321 | 16 | Inverse variance weighted | -0.178 | 0.125 | 0.837 (0.655-1.070) | 0.156 |
| | | MR Egger | -0.329 | 0.586 | 0.719 (0.228-2.269) | 0.583 |
| | | Simple mode | -0.029 | 0.315 | 0.971 (0.524-1.803) | 0.928 |
| | | Weighted median | -0.093 | 0.176 | 0.911 (0.645-1.288) | 0.598 |
| | | Weighted mode | -0.035 | 0.293 | 0.965 (0.543-1.714) | 0.905 |
| Lachnospiraceae UCG004 id.11324 | 13 | Inverse variance weighted | -0.081 | 0.167 | 0.922 (0.665-1.278) | 0.626 |
| | | MR Egger | 0.619 | 0.623 | 1.856 (0.548-6.292) | 0.342 |
| | | Simple mode | 0.105 | 0.380 | 1.111 (0.528-2.337) | 0.787 |
| | | Weighted median | 0.035 | 0.226 | 1.036 (0.665-1.612) | 0.877 |
| | | Weighted mode | 0.105 | 0.342 | 1.111 (0.568-2.171) | 0.764 |
| Lachnospiraceae UCG008 id.11328 | 12 | Inverse variance weighted | -0.039 | 0.139 | 0.962 (0.733-1.262) | 0.778 |
| | | MR Egger | 0.950 | 0.677 | 2.585 (0.686-9.742) | 0.191 |
| | | Simple mode | -0.328 | 0.342 | 0.720 (0.368-1.408) | 0.358 |
| | | Weighted median | -0.159 | 0.177 | 0.853 (0.603-1.207) | 0.369 |
| | | Weighted mode | -0.289 | 0.336 | 0.749 (0.387-1.448) | 0.408 |
| Lachnospiraceae UCG010 id.11330 | 10 | Inverse variance weighted | 0.001 | 0.169 | 1.001 (0.719-1.394) | 0.994 |

| | | | | | | |
|---|---|---|---|---|---|---|
| | | MR Egger | 0.172 | 0.460 | 1.188 (0.482-2.926) | 0.718 |
| | | Simple mode | 0.266 | 0.346 | 1.305 (0.663-2.569) | 0.461 |
| | | Weighted median | 0.015 | 0.223 | 1.015 (0.656-1.572) | 0.945 |
| | | Weighted mode | 0.114 | 0.309 | 1.121 (0.612-2.051) | 0.721 |
| Lachnospira id.2004 | 1 | Wald ratio | -0.422 | 0.644 | 0.656 (0.186-2.316) | 0.512 |
| Lactobacillus id.1837 | 10 | Inverse variance weighted | -0.039 | 0.125 | 0.961 (0.753-1.227) | 0.752 |
| | | MR Egger | -0.322 | 0.361 | 0.724 (0.357-1.470) | 0.398 |
| | | Simple mode | -0.205 | 0.271 | 0.814 (0.479-1.384) | 0.468 |
| | | Weighted median | -0.188 | 0.157 | 0.829 (0.609-1.128) | 0.232 |
| | | Weighted mode | -0.205 | 0.254 | 0.814 (0.495-1.340) | 0.440 |
| Lactococcus id.1851 | 10 | Inverse variance weighted | 0.017 | 0.111 | 1.017 (0.819-1.263) | 0.879 |
| | | MR Egger | -0.629 | 0.556 | 0.533 (0.179-1.586) | 0.291 |
| | | Simple mode | -0.175 | 0.239 | 0.840 (0.526-1.341) | 0.483 |
| | | Weighted median | -0.061 | 0.139 | 0.941 (0.717-1.234) | 0.658 |
| | | Weighted mode | -0.175 | 0.220 | 0.840 (0.545-1.293) | 0.448 |
| Marvinbryantia id.2005 | 10 | Inverse variance weighted | -0.026 | 0.173 | 0.974 (0.694-1.367) | 0.879 |
| | | MR Egger | -0.443 | 0.675 | 0.642 (0.171-2.410) | 0.530 |
| | | Simple mode | -0.042 | 0.353 | 0.959 (0.480-1.914) | 0.907 |
| | | Weighted median | 0.000 | 0.208 | 1.000 (0.665-1.505) | 0.999 |
| | | Weighted mode | -0.024 | 0.361 | 0.977 (0.481-1.981) | 0.949 |
| Methanobrevibacter id.123 | 8 | Inverse variance weighted | 0.298 | 0.130 | 1.347 (1.045-1.736) | 0.022 |
| | | MR Egger | 0.464 | 0.525 | 1.590 (0.568-4.450) | 0.411 |
| | | Simple mode | 0.393 | 0.265 | 1.482 (0.881-2.493) | 0.182 |
| | | Weighted median | 0.233 | 0.154 | 1.262 (0.933-1.707) | 0.131 |
| | | Weighted mode | 0.122 | 0.257 | 1.130 (0.683-1.868) | 0.650 |
| Odoribacter id.952 | 8 | Inverse variance weighted | 0.161 | 0.216 | 1.174 (0.769-1.792) | 0.456 |
| | | MR Egger | -0.353 | 0.700 | 0.703 (0.178-2.770) | 0.632 |
| | | Simple mode | 0.141 | 0.394 | 1.151 (0.532-2.490) | 0.731 |
| | | Weighted median | 0.097 | 0.274 | 1.102 (0.644-1.884) | 0.723 |
| | | Weighted mode | 0.130 | 0.381 | 1.139 (0.539-2.405) | 0.744 |
| Olsenella id.822 | 10 | Inverse variance weighted | -0.008 | 0.141 | 0.992 (0.753-1.307) | 0.955 |
| | | MR Egger | -0.029 | 0.538 | 0.972 (0.338-2.791) | 0.959 |
| | | Simple mode | -0.010 | 0.244 | 0.990 (0.614-1.598) | 0.969 |
| | | Weighted median | -0.045 | 0.141 | 0.956 (0.725-1.260) | 0.748 |
| | | Weighted mode | 0.009 | 0.228 | 1.009 (0.645-1.579) | 0.970 |
| Oscillibacter id.2063 | 15 | Inverse variance weighted | -0.014 | 0.166 | 0.986 (0.713-1.364) | 0.932 |
| | | MR Egger | -0.684 | 0.632 | 0.505 (0.146-1.741) | 0.299 |
| | | Simple mode | -0.426 | 0.382 | 0.653 (0.309-1.381) | 0.284 |
| | | Weighted median | -0.257 | 0.180 | 0.774 (0.543-1.101) | 0.154 |
| | | Weighted mode | -0.434 | 0.372 | 0.648 (0.313-1.344) | 0.263 |
| Oscillospira id.2064 | 9 | Inverse variance weighted | 0.046 | 0.189 | 1.047 (0.723-1.516) | 0.808 |
| | | MR Egger | -0.075 | 0.857 | 0.928 (0.173-4.977) | 0.933 |
| | | Simple mode | -0.097 | 0.301 | 0.907 (0.503-1.637) | 0.755 |
| | | Weighted median | -0.065 | 0.227 | 0.937 (0.601-1.461) | 0.774 |
| | | Weighted mode | -0.097 | 0.297 | 0.907 (0.507-1.624) | 0.751 |
| Oxalobacter id.2978 | 12 | Inverse variance weighted | 0.080 | 0.096 | 1.083 (0.897-1.308) | 0.406 |
| | | MR Egger | -0.482 | 0.413 | 0.617 (0.275-1.387) | 0.270 |
| | | Simple mode | 0.182 | 0.204 | 1.200 (0.804-1.791) | 0.391 |
| | | Weighted median | 0.166 | 0.129 | 1.180 (0.917-1.519) | 0.197 |
| | | Weighted mode | 0.189 | 0.188 | 1.208 (0.837-1.745) | 0.335 |
| Parabacteroides id.954 | 9 | Inverse variance weighted | 0.001 | 0.204 | 1.001 (0.672-1.493) | 0.995 |
| | | MR Egger | -0.108 | 0.562 | 0.898 (0.299-2.699) | 0.853 |

| | | | | | | |
|---|---|---|---|---|---|---|
| | | Simple mode | 0.034 | 0.402 | 1.035 (0.470-2.276) | 0.935 |
| | | Weighted median | -0.018 | 0.267 | 0.982 (0.582-1.657) | 0.946 |
| | | Weighted mode | -0.029 | 0.362 | 0.972 (0.478-1.975) | 0.939 |
| Paraprevotella id.962 | 13 | Inverse variance weighted | -0.003 | 0.126 | 0.997 (0.779-1.276) | 0.983 |
| | | MR Egger | 0.146 | 0.429 | 1.157 (0.499-2.683) | 0.740 |
| | | Simple mode | -0.062 | 0.240 | 0.940 (0.588-1.504) | 0.801 |
| | | Weighted median | -0.072 | 0.149 | 0.930 (0.694-1.247) | 0.629 |
| | | Weighted mode | -0.073 | 0.217 | 0.930 (0.608-1.423) | 0.744 |
| Parasutterella id.2892 | 16 | Inverse variance weighted | 0.029 | 0.126 | 1.030 (0.804-1.318) | 0.816 |
| | | MR Egger | -0.086 | 0.397 | 0.917 (0.421-1.998) | 0.831 |
| | | Simple mode | 0.053 | 0.339 | 1.054 (0.542-2.050) | 0.878 |
| | | Weighted median | 0.024 | 0.179 | 1.024 (0.721-1.455) | 0.893 |
| | | Weighted mode | 0.053 | 0.292 | 1.054 (0.595-1.868) | 0.859 |
| Peptococcus id.2037 | 16 | Inverse variance weighted | 0.102 | 0.101 | 1.107 (0.909-1.350) | 0.313 |
| | | MR Egger | -0.373 | 0.368 | 0.689 (0.335-1.417) | 0.329 |
| | | Simple mode | -0.089 | 0.222 | 0.915 (0.593-1.413) | 0.695 |
| | | Weighted median | -0.015 | 0.128 | 0.985 (0.767-1.264) | 0.904 |
| | | Weighted mode | -0.078 | 0.213 | 0.925 (0.609-1.405) | 0.721 |
| Phascolarctobacterium id.2168 | 10 | Inverse variance weighted | -0.165 | 0.162 | 0.848 (0.618-1.165) | 0.309 |
| | 10 | MR Egger | -1.104 | 0.610 | 0.332 (0.100-1.096) | 0.108 |
| | 10 | Simple mode | -0.212 | 0.332 | 0.809 (0.422-1.550) | 0.539 |
| | 10 | Weighted median | -0.125 | 0.217 | 0.882 (0.576-1.350) | 0.564 |
| | 10 | Weighted mode | -0.217 | 0.323 | 0.805 (0.427-1.518) | 0.519 |
| Prevotella7 id.11182 | 11 | Inverse variance weighted | -0.071 | 0.093 | 0.932 (0.777-1.117) | 0.445 |
| | | MR Egger | -0.339 | 0.558 | 0.713 (0.239-2.129) | 0.559 |
| | | Simple mode | -0.104 | 0.189 | 0.901 (0.622-1.306) | 0.596 |
| | | Weighted median | -0.094 | 0.116 | 0.910 (0.725-1.143) | 0.419 |
| | | Weighted mode | -0.093 | 0.178 | 0.911 (0.642-1.292) | 0.612 |
| Prevotella9 id.11183 | 18 | Inverse variance weighted | 0.139 | 0.110 | 1.149 (0.926-1.427) | 0.207 |
| | | MR Egger | 0.010 | 0.295 | 1.010 (0.566-1.801) | 0.974 |
| | | Simple mode | 0.065 | 0.258 | 1.067 (0.644-1.768) | 0.805 |
| | | Weighted median | 0.110 | 0.147 | 1.117 (0.838-1.488) | 0.452 |
| | | Weighted mode | 0.070 | 0.225 | 1.073 (0.690-1.668) | 0.758 |
| Rikenellaceae RC9 gut group id.11191 | 11 | Inverse variance weighted | -0.125 | 0.110 | 0.882 (0.711-1.095) | 0.255 |
| | | MR Egger | -0.936 | 0.658 | 0.392 (0.108-1.425) | 0.189 |
| | | Simple mode | -0.299 | 0.219 | 0.741 (0.483-1.138) | 0.201 |
| | | Weighted median | -0.143 | 0.129 | 0.866 (0.673-1.115) | 0.265 |
| | | Weighted mode | -0.276 | 0.214 | 0.759 (0.499-1.154) | 0.226 |
| Romboutsia id.11347 | 14 | Inverse variance weighted | 0.138 | 0.144 | 1.148 (0.865-1.523) | 0.340 |
| | | MR Egger | 0.193 | 0.366 | 1.212 (0.592-2.482) | 0.608 |
| | | Simple mode | 0.205 | 0.320 | 1.228 (0.656-2.300) | 0.532 |
| | | Weighted median | 0.134 | 0.194 | 1.143 (0.782-1.671) | 0.490 |
| | | Weighted mode | 0.193 | 0.277 | 1.213 (0.704-2.089) | 0.499 |
| Roseburia id.2012 | 16 | Inverse variance weighted | -0.105 | 0.147 | 0.901 (0.676-1.200) | 0.475 |
| | | MR Egger | 0.131 | 0.333 | 1.140 (0.593-2.188) | 0.701 |
| | | Simple mode | 0.114 | 0.311 | 1.121 (0.610-2.061) | 0.719 |
| | | Weighted median | -0.087 | 0.196 | 0.917 (0.624-1.346) | 0.657 |
| | | Weighted mode | 0.042 | 0.274 | 1.043 (0.610-1.783) | 0.879 |
| Ruminiclostridium5 id.11355 | 14 | Inverse variance weighted | 0.055 | 0.205 | 1.057 (0.707-1.579) | 0.787 |
| | | MR Egger | 0.713 | 0.643 | 2.039 (0.578-7.191) | 0.290 |
| | | Simple mode | 0.496 | 0.545 | 1.643 (0.564-4.782) | 0.379 |

| | | | | | | |
|---|---|---|---|---|---|---|
| | | Weighted median | 0.132 | 0.266 | 1.141 (0.678-1.922) | 0.619 |
| | | Weighted mode | 0.551 | 0.553 | 1.735 (0.587-5.131) | 0.337 |
| Ruminiclostridium6 id.11356 | 15 | Inverse variance weighted | -0.038 | 0.166 | 0.963 (0.696-1.332) | 0.819 |
| | | MR Egger | 0.325 | 0.405 | 1.384 (0.626-3.059) | 0.436 |
| | | Simple mode | 0.090 | 0.396 | 1.094 (0.503-2.379) | 0.824 |
| | | Weighted median | 0.076 | 0.206 | 1.079 (0.721-1.616) | 0.711 |
| | | Weighted mode | 0.129 | 0.356 | 1.138 (0.567-2.284) | 0.722 |
| Ruminiclostridium9 id.11357 | 14 | Inverse variance weighted | -0.007 | 0.172 | 0.993 (0.709-1.390) | 0.966 |
| | | MR Egger | 0.182 | 0.828 | 1.199 (0.237-6.077) | 0.830 |
| | | Simple mode | 0.333 | 0.412 | 1.395 (0.621-3.129) | 0.434 |
| | | Weighted median | 0.110 | 0.235 | 1.116 (0.705-1.767) | 0.640 |
| | | Weighted mode | 0.340 | 0.424 | 1.404 (0.611-3.227) | 0.438 |
| Ruminococcaceae NK4A214 group id.11358 | 14 | Inverse variance weighted | 0.066 | 0.159 | 1.068 (0.782-1.459) | 0.677 |
| | | MR Egger | -0.202 | 0.445 | 0.817 (0.342-1.954) | 0.658 |
| | | Simple mode | 0.438 | 0.375 | 1.550 (0.743-3.235) | 0.264 |
| | | Weighted median | 0.055 | 0.226 | 1.056 (0.678-1.646) | 0.809 |
| | | Weighted mode | 0.218 | 0.344 | 1.244 (0.634-2.441) | 0.536 |
| Ruminococcaceae UCG002 id.11360 | 24 | Inverse variance weighted | 0.016 | 0.138 | 1.016 (0.776-1.331) | 0.908 |
| | | MR Egger | -0.361 | 0.378 | 0.697 (0.333-1.461) | 0.350 |
| | | Simple mode | 0.268 | 0.330 | 1.308 (0.685-2.497) | 0.425 |
| | | Weighted median | 0.004 | 0.181 | 1.004 (0.705-1.430) | 0.982 |
| | | Weighted mode | 0.043 | 0.295 | 1.044 (0.586-1.863) | 0.884 |
| Ruminococcaceae UCG003 id.11361 | 14 | Inverse variance weighted | 0.226 | 0.159 | 1.254 (0.918-1.712) | 0.154 |
| | | MR Egger | 0.432 | 0.544 | 1.541 (0.530-4.479) | 0.442 |
| | | Simple mode | 0.149 | 0.375 | 1.161 (0.557-2.422) | 0.697 |
| | | Weighted median | 0.139 | 0.219 | 1.149 (0.749-1.764) | 0.525 |
| | | Weighted mode | 0.165 | 0.346 | 1.179 (0.599-2.323) | 0.641 |
| Ruminococcaceae UCG004 id.11362 | 10 | Inverse variance weighted | 0.347 | 0.163 | 1.415 (1.029-1.946) | 0.033 |
| | | MR Egger | -0.143 | 0.879 | 0.866 (0.155-4.855) | 0.875 |
| | | Simple mode | 0.648 | 0.377 | 1.912 (0.913-4.006) | 0.120 |
| | | Weighted median | 0.246 | 0.225 | 1.279 (0.823-1.989) | 0.274 |
| | | Weighted mode | 0.059 | 0.338 | 1.061 (0.546-2.060) | 0.865 |
| Ruminococcaceae UCG005 id.11363 | 17 | Inverse variance weighted | -0.234 | 0.141 | 0.792 (0.601-1.043) | 0.097 |
| | | MR Egger | -0.003 | 0.398 | 0.997 (0.457-2.172) | 0.993 |
| | | Simple mode | -0.109 | 0.336 | 0.897 (0.464-1.732) | 0.750 |
| | | Weighted median | -0.131 | 0.187 | 0.877 (0.608-1.266) | 0.484 |
| | | Weighted mode | -0.109 | 0.313 | 0.897 (0.486-1.656) | 0.732 |
| Ruminococcaceae UCG009 id.11366 | 13 | Inverse variance weighted | 0.055 | 0.136 | 1.057 (0.809-1.380) | 0.684 |
| | | MR Egger | 0.154 | 0.595 | 1.167 (0.364-3.742) | 0.800 |
| | | Simple mode | -0.194 | 0.286 | 0.824 (0.470-1.442) | 0.510 |
| | | Weighted median | -0.099 | 0.177 | 0.906 (0.640-1.283) | 0.578 |
| | | Weighted mode | -0.183 | 0.280 | 0.833 (0.481-1.441) | 0.525 |
| Ruminococcaceae UCG010 id.11367 | 7 | Inverse variance weighted | -0.006 | 0.266 | 0.994 (0.591-1.674) | 0.983 |
| | | MR Egger | 0.490 | 1.006 | 1.632 (0.227-11.736) | 0.647 |
| | | Simple mode | 0.178 | 0.491 | 1.195 (0.456-3.131) | 0.729 |

| | | | | | | |
|---|---|---|---|---|---|---|
| | | Weighted median | 0.116 | 0.292 | 1.123 (0.634-1.989) | 0.691 |
| | | Weighted mode | 0.231 | 0.457 | 1.260 (0.514-3.087) | 0.631 |
| Ruminococcaceae UCG011 id.11368 | 8 | Inverse variance weighted | -0.008 | 0.106 | 0.992 (0.805-1.222) | 0.939 |
| | | MR Egger | -0.289 | 0.550 | 0.749 (0.255-2.202) | 0.618 |
| | | Simple mode | -0.070 | 0.200 | 0.933 (0.631-1.380) | 0.738 |
| | | Weighted median | -0.044 | 0.134 | 0.957 (0.736-1.244) | 0.743 |
| | | Weighted mode | -0.072 | 0.199 | 0.930 (0.630-1.373) | 0.727 |
| Ruminococcaceae UCG013 id.11370 | 13 | Inverse variance weighted | -0.127 | 0.169 | 0.881 (0.633-1.226) | 0.452 |
| | | MR Egger | 0.422 | 0.454 | 1.525 (0.626-3.715) | 0.373 |
| | | Simple mode | -0.105 | 0.377 | 0.901 (0.430-1.887) | 0.786 |
| | | Weighted median | 0.071 | 0.239 | 1.073 (0.672-1.714) | 0.767 |
| | | Weighted mode | 0.098 | 0.328 | 1.103 (0.581-2.097) | 0.769 |
| Ruminococcaceae UCG014 id.11371 | 18 | Inverse variance weighted | -0.097 | 0.153 | 0.907 (0.673-1.224) | 0.525 |
| | | MR Egger | -0.121 | 0.476 | 0.886 (0.348-2.255) | 0.803 |
| | | Simple mode | -0.003 | 0.335 | 0.997 (0.517-1.921) | 0.993 |
| | | Weighted median | -0.011 | 0.196 | 0.989 (0.674-1.451) | 0.955 |
| | | Weighted mode | -0.012 | 0.314 | 0.989 (0.535-1.828) | 0.971 |
| Ruminococcus1 id.11373 | 13 | Inverse variance weighted | -0.196 | 0.171 | 0.822 (0.588-1.149) | 0.252 |
| | | MR Egger | -0.269 | 0.474 | 0.764 (0.302-1.934) | 0.581 |
| | | Simple mode | -0.300 | 0.365 | 0.741 (0.362-1.516) | 0.427 |
| | | Weighted median | -0.255 | 0.225 | 0.775 (0.499-1.205) | 0.258 |
| | | Weighted mode | -0.294 | 0.347 | 0.745 (0.377-1.472) | 0.414 |
| Ruminococcus2 id.11374 | 14 | Inverse variance weighted | -0.105 | 0.188 | 0.900 (0.622-1.302) | 0.576 |
| | | MR Egger | -0.474 | 0.508 | 0.622 (0.230-1.684) | 0.369 |
| | | Simple mode | -0.163 | 0.398 | 0.849 (0.389-1.852) | 0.688 |
| | | Weighted median | -0.253 | 0.238 | 0.777 (0.487-1.238) | 0.288 |
| | | Weighted mode | -0.290 | 0.343 | 0.749 (0.382-1.466) | 0.414 |
| Ruminococcus gauvreauii group id.11342 | 13 | Inverse variance weighted | 0.004 | 0.160 | 1.004 (0.733-1.374) | 0.982 |
| | | MR Egger | -0.742 | 0.715 | 0.476 (0.117-1.934) | 0.322 |
| | | Simple mode | -0.096 | 0.348 | 0.908 (0.459-1.797) | 0.787 |
| | | Weighted median | -0.092 | 0.218 | 0.912 (0.595-1.399) | 0.674 |
| | | Weighted mode | -0.096 | 0.365 | 0.908 (0.445-1.856) | 0.797 |
| Ruminococcus gnavus group id.14376 | 12 | Inverse variance weighted | -0.073 | 0.116 | 0.929 (0.741-1.166) | 0.526 |
| | | MR Egger | 0.552 | 0.517 | 1.736 (0.630-4.782) | 0.311 |
| | | Simple mode | -0.156 | 0.261 | 0.856 (0.513-1.429) | 0.564 |
| | | Weighted median | -0.130 | 0.148 | 0.878 (0.657-1.174) | 0.380 |
| | | Weighted mode | -0.184 | 0.241 | 0.832 (0.518-1.334) | 0.461 |
| Ruminococcus torques group id.14377 | 13 | Inverse variance weighted | 0.473 | 0.177 | 1.604 (1.134-2.269) | 0.008 |
| | | MR Egger | 1.324 | 0.523 | 3.757 (1.349-10.465) | 0.028 |
| | | Simple mode | 0.763 | 0.450 | 2.145 (0.888-5.185) | 0.116 |
| | | Weighted median | 0.619 | 0.254 | 1.856 (1.129-3.052) | 0.015 |
| | | Weighted mode | 0.755 | 0.406 | 2.127 (0.961-4.710) | 0.087 |
| Sellimonas id.14369 | 11 | Inverse variance weighted | 0.209 | 0.086 | 1.233 (1.041-1.459) | 0.015 |
| | | MR Egger | 0.099 | 0.515 | 1.104 (0.403-3.026) | 0.852 |
| | | Simple mode | 0.194 | 0.176 | 1.214 (0.860-1.716) | 0.296 |
| | | Weighted median | 0.186 | 0.115 | 1.205 (0.962-1.509) | 0.105 |

| | | | | | | |
|---|---|---|---|---|---|---|
| | | Weighted mode | 0.183 | 0.172 | 1.201 (0.857-1.683) | 0.313 |
| Senegalimassilia id.11160 | 5 | Inverse variance weighted | -0.099 | 0.183 | 0.905 (0.632-1.297) | 0.588 |
| | | MR Egger | 0.016 | 0.646 | 1.016 (0.286-3.608) | 0.982 |
| | | Simple mode | -0.054 | 0.298 | 0.947 (0.528-1.699) | 0.864 |
| | | Weighted median | -0.037 | 0.238 | 0.964 (0.605-1.535) | 0.876 |
| | | Weighted mode | -0.051 | 0.272 | 0.950 (0.558-1.619) | 0.860 |
| Slackia id.825 | 9 | Inverse variance weighted | 0.032 | 0.135 | 1.032 (0.793-1.344) | 0.814 |
| | | MR Egger | 0.534 | 0.579 | 1.705 (0.549-5.301) | 0.387 |
| | | Simple mode | 0.323 | 0.338 | 1.382 (0.712-2.681) | 0.367 |
| | | Weighted median | 0.105 | 0.188 | 1.111 (0.769-1.606) | 0.575 |
| | | Weighted mode | 0.337 | 0.349 | 1.401 (0.707-2.777) | 0.362 |
| Streptococcus id.1853 | 16 | Inverse variance weighted | 0.064 | 0.165 | 1.066 (0.771-1.475) | 0.698 |
| | | MR Egger | 0.871 | 0.616 | 2.390 (0.715-7.992) | 0.179 |
| | | Simple mode | -0.327 | 0.443 | 0.721 (0.303-1.719) | 0.472 |
| | | Weighted median | -0.101 | 0.227 | 0.904 (0.579-1.409) | 0.655 |
| | | Weighted mode | -0.302 | 0.476 | 0.739 (0.291-1.878) | 0.535 |
| Subdoligranulum id.2070 | 13 | Inverse variance weighted | 0.224 | 0.179 | 1.251 (0.882-1.775) | 0.210 |
| | | MR Egger | 0.204 | 0.491 | 1.227 (0.468-3.214) | 0.685 |
| | | Simple mode | 0.394 | 0.400 | 1.483 (0.677-3.245) | 0.344 |
| | | Weighted median | 0.314 | 0.247 | 1.369 (0.845-2.220) | 0.202 |
| | | Weighted mode | 0.370 | 0.390 | 1.448 (0.674-3.112) | 0.361 |
| Sutterella id.2896 | 12 | Inverse variance weighted | 0.080 | 0.177 | 1.084 (0.766-1.534) | 0.650 |
| | | MR Egger | 0.356 | 0.870 | 1.427 (0.259-7.858) | 0.691 |
| | | Simple mode | -0.274 | 0.364 | 0.760 (0.373-1.551) | 0.467 |
| | | Weighted median | -0.044 | 0.232 | 0.957 (0.608-1.506) | 0.848 |
| | | Weighted mode | -0.268 | 0.354 | 0.765 (0.382-1.532) | 0.465 |
| Terrisporobacter id.11348 | 5 | Inverse variance weighted | 0.360 | 0.176 | 1.434 (1.016-2.023) | 0.040 |
| | | MR Egger | 0.029 | 0.481 | 1.029 (0.401-2.644) | 0.956 |
| | | Simple mode | 0.268 | 0.283 | 1.307 (0.750-2.276) | 0.398 |
| | | Weighted median | 0.272 | 0.221 | 1.312 (0.850-2.024) | 0.220 |
| | | Weighted mode | 0.270 | 0.296 | 1.310 (0.734-2.338) | 0.413 |
| Turicibacter id.2162 | 13 | Inverse variance weighted | -0.159 | 0.136 | 0.853 (0.653-1.115) | 0.245 |
| | | MR Egger | -0.127 | 0.597 | 0.881 (0.273-2.837) | 0.835 |
| | | Simple mode | 0.088 | 0.323 | 1.092 (0.579-2.058) | 0.790 |
| | | Weighted median | -0.029 | 0.187 | 0.972 (0.673-1.402) | 0.878 |
| | | Weighted mode | 0.075 | 0.316 | 1.078 (0.580-2.003) | 0.816 |
| Tyzzerella3 id.11335 | 14 | Inverse variance weighted | 0.036 | 0.102 | 1.037 (0.849-1.266) | 0.723 |
| | | MR Egger | -0.526 | 0.581 | 0.591 (0.189-1.847) | 0.384 |
| | | Simple mode | 0.029 | 0.272 | 1.029 (0.604-1.754) | 0.918 |
| | | Weighted median | 0.032 | 0.137 | 1.032 (0.790-1.349) | 0.816 |
| | | Weighted mode | 0.224 | 0.239 | 1.251 (0.783-1.998) | 0.366 |
| unknown genus id.1000000073 | 15 | Inverse variance weighted | -0.081 | 0.154 | 0.923 (0.683-1.247) | 0.600 |
| | | MR Egger | -0.051 | 0.449 | 0.950 (0.394-2.290) | 0.911 |
| | | Simple mode | -0.335 | 0.310 | 0.716 (0.390-1.315) | 0.299 |
| | | Weighted median | -0.215 | 0.179 | 0.806 (0.568-1.146) | 0.230 |
| | | Weighted mode | -0.310 | 0.287 | 0.733 (0.418-1.287) | 0.298 |
| unknown genus id.1000001215 | 11 | Inverse variance weighted | -0.179 | 0.112 | 0.836 (0.671-1.043) | 0.112 |
| | | MR Egger | -0.056 | 0.327 | 0.946 (0.498-1.797) | 0.868 |
| | | Simple mode | -0.446 | 0.265 | 0.640 (0.381-1.076) | 0.123 |
| | | Weighted median | -0.227 | 0.153 | 0.797 (0.590-1.075) | 0.137 |
| | | Weighted mode | -0.433 | 0.249 | 0.649 (0.398-1.056) | 0.112 |
| unknown genus id.1000005472 | 14 | Inverse variance weighted | 0.179 | 0.137 | 1.196 (0.915-1.564) | 0.190 |

| | | | | | | |
|---|---|---|---|---|---|---|
| | | MR Egger | 0.107 | 0.420 | 1.113 (0.489-2.534) | 0.803 |
| | | Simple mode | 0.350 | 0.316 | 1.418 (0.763-2.637) | 0.289 |
| | | Weighted median | 0.253 | 0.183 | 1.288 (0.899-1.845) | 0.167 |
| | | Weighted mode | 0.350 | 0.291 | 1.418 (0.803-2.507) | 0.250 |
| unknown genus id.1000005479 | 10 | Inverse variance weighted | 0.140 | 0.206 | 1.150 (0.768-1.723) | 0.498 |
| | | MR Egger | 0.512 | 1.072 | 1.669 (0.204-13.651) | 0.646 |
| | | Simple mode | 0.704 | 0.459 | 2.021 (0.823-4.968) | 0.159 |
| | | Weighted median | 0.046 | 0.220 | 1.047 (0.680-1.613) | 0.833 |
| | | Weighted mode | 0.642 | 0.434 | 1.900 (0.811-4.450) | 0.173 |
| unknown genus id.1000006162 | 15 | Inverse variance weighted | -0.037 | 0.092 | 0.964 (0.805-1.154) | 0.687 |
| | | MR Egger | -0.215 | 0.405 | 0.806 (0.365-1.784) | 0.604 |
| | | Simple mode | 0.053 | 0.232 | 1.054 (0.669-1.661) | 0.824 |
| | | Weighted median | 0.006 | 0.119 | 1.006 (0.797-1.270) | 0.959 |
| | | Weighted mode | 0.053 | 0.218 | 1.054 (0.688-1.616) | 0.813 |
| unknown genus id.1868 | 13 | Inverse variance weighted | -0.063 | 0.133 | 0.939 (0.724-1.218) | 0.633 |
| | | MR Egger | 0.132 | 0.401 | 1.141 (0.519-2.506) | 0.749 |
| | | Simple mode | -0.359 | 0.298 | 0.699 (0.390-1.252) | 0.251 |
| | | Weighted median | -0.203 | 0.180 | 0.816 (0.574-1.160) | 0.258 |
| | | Weighted mode | -0.348 | 0.338 | 0.706 (0.364-1.368) | 0.322 |
| unknown genus id.2001 | 11 | Inverse variance weighted | -0.011 | 0.182 | 0.989 (0.692-1.414) | 0.951 |
| | | MR Egger | -0.111 | 0.649 | 0.895 (0.251-3.193) | 0.868 |
| | | Simple mode | -0.112 | 0.344 | 0.894 (0.456-1.756) | 0.752 |
| | | Weighted median | -0.023 | 0.207 | 0.977 (0.652-1.465) | 0.911 |
| | | Weighted mode | -0.060 | 0.335 | 0.941 (0.488-1.816) | 0.860 |
| unknown genus id.2041 | 12 | Inverse variance weighted | -0.027 | 0.126 | 0.973 (0.760-1.245) | 0.829 |
| | | MR Egger | 0.644 | 0.375 | 1.903 (0.913-3.969) | 0.117 |
| | | Simple mode | -0.184 | 0.299 | 0.832 (0.463-1.495) | 0.551 |
| | | Weighted median | 0.003 | 0.174 | 1.003 (0.712-1.411) | 0.988 |
| | | Weighted mode | 0.031 | 0.285 | 1.032 (0.590-1.805) | 0.915 |
| unknown genus id.2071 | 18 | Inverse variance weighted | 0.081 | 0.185 | 1.085 (0.755-1.559) | 0.660 |
| | | MR Egger | 2.426 | 0.749 | 11.314 (2.606-49.110) | 0.005 |
| | | Simple mode | 0.775 | 0.498 | 2.170 (0.818-5.760) | 0.138 |
| | | Weighted median | 0.072 | 0.218 | 1.075 (0.701-1.647) | 0.741 |
| | | Weighted mode | -0.419 | 0.472 | 0.657 (0.261-1.659) | 0.387 |
| unknown genus id.2755 | 15 | Inverse variance weighted | 0.135 | 0.138 | 1.145 (0.874-1.500) | 0.327 |
| | | MR Egger | -0.371 | 0.563 | 0.690 (0.229-2.082) | 0.522 |
| | | Simple mode | 0.309 | 0.347 | 1.362 (0.690-2.687) | 0.388 |
| | | Weighted median | 0.158 | 0.177 | 1.171 (0.828-1.657) | 0.371 |
| | | Weighted mode | 0.275 | 0.319 | 1.316 (0.705-2.458) | 0.403 |
| unknown genus id.826 | 15 | Inverse variance weighted | -0.184 | 0.176 | 0.832 (0.590-1.174) | 0.294 |
| | | MR Egger | 0.391 | 0.459 | 1.479 (0.602-3.632) | 0.409 |
| | | Simple mode | 0.287 | 0.435 | 1.332 (0.568-3.126) | 0.520 |
| | | Weighted median | -0.067 | 0.225 | 0.935 (0.602-1.454) | 0.766 |
| | | Weighted mode | 0.154 | 0.307 | 1.166 (0.639-2.128) | 0.624 |
| unknown genus id.959 | 13 | Inverse variance weighted | 0.052 | 0.098 | 1.053 (0.869-1.276) | 0.596 |
| | | MR Egger | 0.272 | 0.631 | 1.312 (0.381-4.517) | 0.675 |
| | | Simple mode | 0.105 | 0.211 | 1.111 (0.735-1.680) | 0.626 |
| | | Weighted median | 0.077 | 0.131 | 1.080 (0.835-1.398) | 0.556 |
| | | Weighted mode | 0.118 | 0.211 | 1.125 (0.744-1.703) | 0.586 |
| Veillonella id.2198 | 10 | Inverse variance weighted | -0.011 | 0.149 | 0.989 (0.738-1.325) | 0.939 |
| | | MR Egger | -1.315 | 0.798 | 0.269 (0.056-1.284) | 0.138 |
| | | Simple mode | 0.183 | 0.329 | 1.201 (0.630-2.289) | 0.591 |

| | | | | | | | |
|---|---|---|---|---|---|---|---|
| | | | Weighted median | 0.098 | 0.203 | 1.103 (0.741-1.640) | 0.630 |
| | | | Weighted mode | 0.174 | 0.353 | 1.189 (0.596-2.374) | 0.634 |
| | Victivallis id.2256 | 12 | Inverse variance weighted | -0.120 | 0.102 | 0.887 (0.726-1.084) | 0.241 |
| | | | MR Egger | -1.105 | 0.619 | 0.331 (0.099-1.114) | 0.105 |
| | | | Simple mode | -0.122 | 0.181 | 0.885 (0.621-1.260) | 0.511 |
| | | | Weighted median | -0.128 | 0.112 | 0.880 (0.707-1.096) | 0.255 |
| | | | Weighted mode | -0.122 | 0.172 | 0.885 (0.632-1.239) | 0.491 |
| order | Actinomycetales id.420 | 5 | Inverse variance weighted | 0.184 | 0.185 | 1.202 (0.837-1.725) | 0.320 |
| | | | MR Egger | 0.152 | 0.473 | 1.164 (0.461-2.943) | 0.769 |
| | | | Simple mode | 0.233 | 0.296 | 1.263 (0.707-2.256) | 0.475 |
| | | | Weighted median | 0.241 | 0.219 | 1.273 (0.829-1.955) | 0.270 |
| | | | Weighted mode | 0.266 | 0.290 | 1.305 (0.740-2.303) | 0.410 |
| | Bacillales id.1674 | 11 | Inverse variance weighted | -0.080 | 0.111 | 0.924 (0.743-1.148) | 0.473 |
| | | | MR Egger | 0.325 | 0.529 | 1.384 (0.491-3.904) | 0.554 |
| | | | Simple mode | 0.153 | 0.239 | 1.165 (0.729-1.863) | 0.537 |
| | | | Weighted median | 0.018 | 0.131 | 1.019 (0.788-1.317) | 0.889 |
| | | | Weighted mode | 0.136 | 0.226 | 1.145 (0.735-1.784) | 0.562 |
| | Bacteroidales id.913 | 17 | Inverse variance weighted | 0.448 | 0.152 | 1.565 (1.161-2.108) | 0.003 |
| | | | MR Egger | 0.186 | 0.326 | 1.204 (0.635-2.283) | 0.577 |
| | | | Simple mode | 0.252 | 0.326 | 1.287 (0.679-2.438) | 0.451 |
| | | | Weighted median | 0.416 | 0.224 | 1.516 (0.977-2.353) | 0.063 |
| | | | Weighted mode | 0.358 | 0.296 | 1.431 (0.800-2.558) | 0.244 |
| | Bifidobacteriales id.432 | 17 | Inverse variance weighted | 0.072 | 0.147 | 1.075 (0.805-1.435) | 0.624 |
| | | | MR Egger | 0.735 | 0.547 | 2.085 (0.713-6.098) | 0.199 |
| | | | Simple mode | 0.354 | 0.389 | 1.425 (0.665-3.055) | 0.376 |
| | | | Weighted median | 0.188 | 0.215 | 1.207 (0.793-1.839) | 0.380 |
| | | | Weighted mode | 0.224 | 0.309 | 1.251 (0.682-2.294) | 0.479 |
| | Burkholderiales id.2874 | 13 | Inverse variance weighted | -0.121 | 0.177 | 0.886 (0.626-1.254) | 0.494 |
| | | | MR Egger | 0.110 | 0.551 | 1.116 (0.379-3.287) | 0.845 |
| | | | Simple mode | -0.318 | 0.384 | 0.728 (0.343-1.544) | 0.424 |
| | | | Weighted median | -0.270 | 0.229 | 0.763 (0.488-1.195) | 0.238 |
| | | | Weighted mode | -0.301 | 0.393 | 0.740 (0.343-1.598) | 0.458 |
| | Clostridiales id.1863 | 18 | Inverse variance weighted | 0.115 | 0.149 | 1.122 (0.838-1.501) | 0.440 |
| | | | MR Egger | 0.125 | 0.357 | 1.133 (0.563-2.284) | 0.731 |
| | | | Simple mode | 0.113 | 0.341 | 1.120 (0.574-2.184) | 0.743 |
| | | | Weighted median | 0.206 | 0.210 | 1.229 (0.814-1.855) | 0.326 |
| | | | Weighted mode | 0.203 | 0.263 | 1.225 (0.731-2.051) | 0.452 |
| | Coriobacteriales id.810 | 15 | Inverse variance weighted | 0.061 | 0.154 | 1.063 (0.786-1.438) | 0.693 |
| | | | MR Egger | 0.379 | 0.354 | 1.460 (0.730-2.920) | 0.304 |
| | | | Simple mode | 0.493 | 0.379 | 1.638 (0.780-3.439) | 0.214 |
| | | | Weighted median | 0.077 | 0.231 | 1.080 (0.686-1.699) | 0.740 |
| | | | Weighted mode | 0.078 | 0.285 | 1.081 (0.618-1.889) | 0.789 |
| | Desulfovibrionales id.3156 | 13 | Inverse variance weighted | 0.111 | 0.173 | 1.117 (0.796-1.568) | 0.522 |
| | | | MR Egger | -0.096 | 0.493 | 0.909 (0.346-2.386) | 0.849 |
| | | | Simple mode | -0.104 | 0.371 | 0.901 (0.435-1.864) | 0.783 |
| | | | Weighted median | 0.031 | 0.232 | 1.032 (0.655-1.625) | 0.892 |
| | | | Weighted mode | -0.018 | 0.306 | 0.983 (0.539-1.791) | 0.955 |
| | Enterobacteriales id.3468 | 10 | Inverse variance weighted | -0.016 | 0.213 | 0.984 (0.648-1.495) | 0.940 |
| | | | MR Egger | -1.537 | 0.834 | 0.215 (0.042-1.104) | 0.103 |
| | | | Simple mode | 0.055 | 0.496 | 1.056 (0.400-2.790) | 0.915 |
| | | | Weighted median | 0.055 | 0.262 | 1.057 (0.632-1.766) | 0.834 |
| | | | Weighted mode | 0.091 | 0.437 | 1.095 (0.465-2.578) | 0.840 |

| | | | | | | |
|---|---|---|---|---|---|---|
| Erysipelotrichales id.2148 | 13 | Inverse variance weighted | -0.142 | 0.187 | 0.867 (0.601-1.251) | 0.447 |
| | | MR Egger | -1.870 | 0.794 | 0.154 (0.032-0.731) | 0.038 |
| | | Simple mode | -0.296 | 0.424 | 0.744 (0.324-1.708) | 0.498 |
| | | Weighted median | -0.321 | 0.260 | 0.725 (0.436-1.206) | 0.216 |
| | | Weighted mode | -0.289 | 0.414 | 0.749 (0.333-1.687) | 0.499 |
| Gastranaerophilales id.1591 | 11 | Inverse variance weighted | -0.179 | 0.112 | 0.836 (0.671-1.043) | 0.112 |
| | | MR Egger | -0.056 | 0.327 | 0.946 (0.498-1.797) | 0.868 |
| | | Simple mode | -0.446 | 0.279 | 0.640 (0.371-1.106) | 0.141 |
| | | Weighted median | -0.227 | 0.153 | 0.797 (0.590-1.076) | 0.138 |
| | | Weighted mode | -0.433 | 0.246 | 0.649 (0.400-1.051) | 0.109 |
| Lactobacillales id.1800 | 19 | Inverse variance weighted | 0.086 | 0.136 | 1.090 (0.835-1.424) | 0.525 |
| | | MR Egger | 0.386 | 0.336 | 1.470 (0.760-2.843) | 0.268 |
| | | Simple mode | 0.042 | 0.347 | 1.043 (0.529-2.058) | 0.904 |
| | | Weighted median | 0.047 | 0.184 | 1.048 (0.731-1.503) | 0.798 |
| | | Weighted mode | 0.028 | 0.285 | 1.028 (0.588-1.799) | 0.923 |
| Methanobacteriales id.120 | 12 | Inverse variance weighted | 0.092 | 0.138 | 1.096 (0.836-1.437) | 0.507 |
| | | MR Egger | 0.210 | 0.562 | 1.233 (0.410-3.710) | 0.717 |
| | | Simple mode | 0.205 | 0.271 | 1.227 (0.722-2.087) | 0.465 |
| | | Weighted median | 0.074 | 0.137 | 1.077 (0.824-1.409) | 0.587 |
| | | Weighted mode | -0.001 | 0.269 | 0.999 (0.590-1.692) | 0.997 |
| Mollicutes RF9 id.11579 | 14 | Inverse variance weighted | 0.179 | 0.137 | 1.196 (0.915-1.564) | 0.190 |
| | | MR Egger | 0.107 | 0.420 | 1.113 (0.489-2.534) | 0.803 |
| | | Simple mode | 0.350 | 0.300 | 1.418 (0.788-2.553) | 0.265 |
| | | Weighted median | 0.253 | 0.187 | 1.288 (0.894-1.857) | 0.175 |
| | | Weighted mode | 0.350 | 0.294 | 1.418 (0.797-2.523) | 0.256 |
| NB1n id.3953 | 15 | Inverse variance weighted | -0.037 | 0.092 | 0.964 (0.805-1.154) | 0.687 |
| | | MR Egger | -0.215 | 0.405 | 0.806 (0.365-1.784) | 0.604 |
| | | Simple mode | 0.053 | 0.210 | 1.054 (0.699-1.590) | 0.806 |
| | | Weighted median | 0.006 | 0.125 | 1.006 (0.788-1.285) | 0.961 |
| | | Weighted mode | 0.053 | 0.214 | 1.054 (0.693-1.603) | 0.809 |
| Pasteurellales id.3688 | 17 | Inverse variance weighted | 0.112 | 0.106 | 1.119 (0.909-1.376) | 0.290 |
| | | MR Egger | 0.140 | 0.261 | 1.150 (0.689-1.918) | 0.601 |
| | | Simple mode | 0.299 | 0.263 | 1.349 (0.806-2.258) | 0.271 |
| | | Weighted median | 0.153 | 0.148 | 1.165 (0.872-1.556) | 0.301 |
| | | Weighted mode | 0.279 | 0.225 | 1.321 (0.850-2.054) | 0.234 |
| Rhodospirillales id.2667 | 15 | Inverse variance weighted | 0.216 | 0.122 | 1.241 (0.977-1.575) | 0.076 |
| | | MR Egger | -0.763 | 0.549 | 0.466 (0.159-1.367) | 0.188 |
| | | Simple mode | 0.373 | 0.309 | 1.452 (0.792-2.659) | 0.248 |
| | | Weighted median | 0.218 | 0.169 | 1.243 (0.893-1.731) | 0.197 |
| | | Weighted mode | 0.367 | 0.302 | 1.443 (0.799-2.607) | 0.244 |
| Selenomonadales id.2165 | 13 | Inverse variance weighted | 0.087 | 0.181 | 1.091 (0.764-1.557) | 0.631 |
| | | MR Egger | -0.166 | 0.584 | 0.847 (0.270-2.659) | 0.781 |
| | | Simple mode | -0.208 | 0.424 | 0.812 (0.354-1.864) | 0.633 |
| | | Weighted median | -0.094 | 0.244 | 0.910 (0.564-1.468) | 0.699 |
| | | Weighted mode | -0.201 | 0.398 | 0.818 (0.375-1.786) | 0.623 |
| Verrucomicrobiales id.4030 | 12 | Inverse variance weighted | 0.198 | 0.196 | 1.219 (0.830-1.790) | 0.313 |
| | | MR Egger | -0.795 | 0.659 | 0.452 (0.124-1.645) | 0.256 |
| | | Simple mode | 0.183 | 0.427 | 1.201 (0.519-2.776) | 0.677 |
| | | Weighted median | 0.219 | 0.222 | 1.245 (0.807-1.923) | 0.322 |
| | | Weighted mode | 0.148 | 0.413 | 1.159 (0.516-2.602) | 0.727 |
| Victivallales id.2254 | 10 | Inverse variance weighted | -0.286 | 0.125 | 0.751 (0.588-0.960) | 0.022 |
| | | MR Egger | -0.517 | 0.475 | 0.596 (0.235-1.511) | 0.307 |

| | | | | | | | |
|---|---|---|---|---|---|---|---|
| | | | Simple mode | -0.072 | 0.268 | 0.930 (0.550-1.573) | 0.794 |
| | | | Weighted median | -0.130 | 0.152 | 0.878 (0.652-1.182) | 0.391 |
| | | | Weighted mode | -0.072 | 0.241 | 0.930 (0.580-1.492) | 0.772 |
| phylum | Actinobacteria id.400 | 16 | Inverse variance weighted | -0.063 | 0.160 | 0.939 (0.687-1.284) | 0.693 |
| | | | MR Egger | -0.236 | 0.667 | 0.790 (0.214-2.920) | 0.729 |
| | | | Simple mode | 0.075 | 0.418 | 1.078 (0.475-2.446) | 0.860 |
| | | | Weighted median | -0.003 | 0.225 | 0.997 (0.642-1.550) | 0.991 |
| | | | Weighted mode | 0.124 | 0.375 | 1.132 (0.543-2.359) | 0.746 |
| | Bacteroidetes id.905 | 14 | Inverse variance weighted | 0.266 | 0.190 | 1.304 (0.898-1.894) | 0.163 |
| | | | MR Egger | 0.380 | 0.436 | 1.462 (0.621-3.439) | 0.402 |
| | | | Simple mode | 0.156 | 0.376 | 1.169 (0.559-2.444) | 0.685 |
| | | | Weighted median | 0.306 | 0.242 | 1.359 (0.845-2.184) | 0.206 |
| | | | Weighted mode | 0.324 | 0.289 | 1.383 (0.784-2.438) | 0.283 |
| | Cyanobacteria id.1500 | 10 | Inverse variance weighted | 0.028 | 0.126 | 1.029 (0.803-1.318) | 0.823 |
| | | | MR Egger | -0.506 | 0.405 | 0.603 (0.273-1.333) | 0.247 |
| | | | Simple mode | 0.068 | 0.235 | 1.070 (0.675-1.698) | 0.779 |
| | | | Weighted median | 0.031 | 0.166 | 1.032 (0.745-1.428) | 0.851 |
| | | | Weighted mode | 0.053 | 0.253 | 1.055 (0.642-1.732) | 0.838 |
| | Euryarchaeota id.55 | 13 | Inverse variance weighted | 0.166 | 0.105 | 1.181 (0.962-1.450) | 0.113 |
| | | | MR Egger | 1.324 | 0.366 | 3.759 (1.834-7.704) | 0.004 |
| | | | Simple mode | -0.021 | 0.240 | 0.979 (0.611-1.568) | 0.932 |
| | | | Weighted median | 0.032 | 0.130 | 1.033 (0.801-1.332) | 0.804 |
| | | | Weighted mode | -0.031 | 0.235 | 0.970 (0.612-1.536) | 0.898 |
| | Firmicutes id.1672 | 21 | Inverse variance weighted | -0.009 | 0.134 | 0.991 (0.762-1.289) | 0.947 |
| | | | MR Egger | 0.400 | 0.306 | 1.491 (0.818-2.719) | 0.208 |
| | | | Simple mode | -0.205 | 0.322 | 0.815 (0.433-1.531) | 0.531 |
| | | | Weighted median | -0.058 | 0.189 | 0.943 (0.651-1.367) | 0.758 |
| | | | Weighted mode | 0.142 | 0.261 | 1.152 (0.691-1.922) | 0.593 |
| | Lentisphaerae id.2238 | 11 | Inverse variance weighted | -0.242 | 0.123 | 0.785 (0.617-0.999) | 0.049 |
| | | | MR Egger | -0.420 | 0.490 | 0.657 (0.251-1.717) | 0.414 |
| | | | Simple mode | -0.003 | 0.243 | 0.997 (0.619-1.605) | 0.990 |
| | | | Weighted median | -0.070 | 0.144 | 0.933 (0.703-1.237) | 0.628 |
| | | | Weighted mode | -0.030 | 0.230 | 0.970 (0.618-1.525) | 0.899 |
| | Proteobacteria id.2375 | 14 | Inverse variance weighted | -0.107 | 0.199 | 0.898 (0.608-1.327) | 0.590 |
| | | | MR Egger | 0.755 | 0.590 | 2.127 (0.669-6.765) | 0.225 |
| | | | Simple mode | 0.068 | 0.394 | 1.071 (0.494-2.320) | 0.865 |
| | | | Weighted median | 0.049 | 0.245 | 1.051 (0.650-1.699) | 0.841 |
| | | | Weighted mode | 0.077 | 0.356 | 1.080 (0.538-2.169) | 0.831 |
| | Tenericutes id.3919 | 12 | Inverse variance weighted | 0.270 | 0.151 | 1.310 (0.975-1.762) | 0.074 |
| | | | MR Egger | -0.161 | 0.487 | 0.851 (0.327-2.213) | 0.748 |
| | | | Simple mode | 0.368 | 0.345 | 1.444 (0.735-2.838) | 0.309 |
| | | | Weighted median | 0.308 | 0.197 | 1.361 (0.924-2.003) | 0.118 |
| | | | Weighted mode | 0.326 | 0.304 | 1.386 (0.764-2.514) | 0.306 |
| | Verrucomicrobia id.3982 | 12 | Inverse variance weighted | 0.122 | 0.165 | 1.130 (0.817-1.563) | 0.460 |
| | | | MR Egger | -0.386 | 0.447 | 0.680 (0.283-1.633) | 0.408 |
| | | | Simple mode | 0.122 | 0.385 | 1.130 (0.532-2.401) | 0.757 |
| | | | Weighted median | 0.083 | 0.214 | 1.086 (0.713-1.653) | 0.700 |
| | | | Weighted mode | 0.085 | 0.362 | 1.088 (0.536-2.210) | 0.819 |

MR, Mendelian randomization；SNPs, single nucleotide polymorphisms; Beta, the effect size of the exposure on 28-day sepsis mortaliry; SE, standard error; OR, odds ratio; CI, confidence interval.
